# Neural pathway activation in the subthalamic region depends on stimulation polarity

**DOI:** 10.1101/2024.05.01.24306044

**Authors:** Seyyed Bahram Borgheai, Enrico Opri, Faical Isbaine, Eric Cole, Roohollah Jafari Deligani, Nealen Laxpati, Benjamin B Risk, Jon T Willie, Robert E Gross, Nicholas Au Yong, Cameron C McIntyre, Svjetlana Miocinovic

**Affiliations:** Department of Neurology, Emory University, Atlanta, GA; Department of Biomedical Engineering, University of Michigan, Ann Arbor, MI; Department of Neurosurgery, Emory University, Atlanta, GA; Department of Biomedical Engineering, Emory University and Georgia Institute of Technology; Department of Biostatistics and Bioinformatics, Emory University, Atlanta, GA; Department of Neurosurgery, Washington University, St Louis, MO; Department of Neurosurgery, Rutgers Robert Wood Johnson medical School, New Brunswick NJ; Department of Cell Biology, Emory University, Atlanta, GA; Department of Biomedical Engineering, Duke University, Durham, NC

## Abstract

Deep brain stimulation (DBS) is an effective treatment for Parkinson’s disease (PD); however, there is limited understanding of which subthalamic pathways are recruited in response to stimulation. Here, by focusing on the polarity of the stimulus waveform (cathodic vs. anodic), our goal was to elucidate biophysical mechanisms that underlie electrical stimulation in the human brain. In clinical studies, cathodic stimulation more easily triggers behavioral responses, but anodic DBS broadens the therapeutic window. This suggests that neural pathways involved respond preferentially depending on stimulus polarity. To experimentally compare the activation of therapeutically relevant pathways during cathodic and anodic subthalamic nucleus (STN) DBS, pathway activation was quantified by measuring evoked potentials resulting from antidromic or orthodromic activation in 15 PD patients undergoing DBS implantation. Cortical evoked potentials (cEP) were recorded using subdural electrocorticography, DBS local evoked potentials (DLEP) were recorded from non-stimulating contacts and EMG activity was recorded from arm and face muscles. We measured: 1) the amplitude of short-latency cEP, previously demonstrated to reflect activation of the cortico-STN hyperdirect pathway, 2) DLEP amplitude thought to reflect activation of STN-globus pallidus (GP) pathway, and 3) amplitudes of very short-latency cEP and motor evoked potentials (mEP) for activation of cortico-spinal/bulbar tract (CSBT). We constructed recruitment and strength-duration curves for each EP/pathway to compare the excitability for different stimulation polarities. We compared experimental data with the most advanced DBS computational models. Our results provide experimental evidence that subcortical cathodic and anodic stimulation activate the same pathways in the STN region and that cathodic stimulation is in general more efficient. However, relative efficiency varies for different pathways so that anodic stimulation is the least efficient in activating CSBT, more efficient in activating the HDP and as efficient as cathodic in activating STN-GP pathway. Our experiments confirm biophysical model predictions regarding neural activations in the central nervous system and provide evidence that stimulus polarity has differential effects on passing axons, terminal synapses, and local neurons. Comparison of experimental results with clinical DBS studies provides further evidence that the hyperdirect pathway may be involved in the therapeutic mechanisms of DBS.

## Introduction

Selective activation of neurons with electrical stimulation is desirable in clinical neuromodulation applications where the aim is to maximize target engagement while limiting off-target effects, but this is difficult to achieve. Despite the effectiveness and relative safety of deep brain stimulation (DBS) therapy for medication-refractory Parkinson’s disease (PD) being established in multiple clinical trials ^1–3^, how stimulation parameters impact recruitment of clinically relevant pathways in the subthalamic region is still not well understood.

In particular, little is known about the effect of stimulus waveform polarity, i.e. cathodic vs. anodic, or their underlying neurophysiological actions. First-generation DBS devices were restricted to cathodic pulses during monopolar stimulation by always utilizing the implanted pulse generator case as the anode. This configuration was supported by clinical and pre-clinical data demonstrating that monopolar cathodic stimulation elicits behavioral responses more readily than anodic stimulation ^4,5^. However, recent clinical studies have demonstrated that anodic DBS can widen the therapeutic window, a difference in a stimulation parameter value that results in therapeutic benefit versus unwanted side effect ^6,7^. Putative pathways responsible for DBS therapeutic benefit include the subthalamopallidal (STN-GP) projections ^8^ and cortico-subthalamic hyperdirect pathway (HDP)^9,10^, while motor side effects (unwanted muscle contractions) are due to activation of the corticospinal and corticobulbar tracts (CSBT) ^11^. Computational studies have suggested that anodic DBS may be more efficient (lower activation threshold) than cathodic when activating terminating axons of the HDP compared to passing axons of the CSBT ^12–14^, however relative activation thresholds also depend on neuronal morphology ^15^. When coupled with fundamental biophysical knowledge, differential responses to cathodic and anodic stimulation can provide novel insights into which neuronal elements are activated by DBS.

Experimental investigations have long sought to dissect the activation of DBS-relevant pathways, albeit almost exclusively with traditional cathodic stimulation. The short latency (2-10 ms) cortical evoked potentials (cEP) recorded over the sensorimotor and prefrontal regions in response to single STN DBS pulses have been characterized as markers of antidromic activation of the HDP ^16–23^. A previous intraoperative study identified three short-latency evoked potentials (EP1, EP2, EP3) over the sensorimotor cortex as related to antidromic HDP activation, with EP1 being related to therapeutic stimulation ^17^. The very short-latency cortical potentials (<2 ms; EP0) have been associated with CSBT activation and accompanied by motor evoked potentials (mEP) recorded peripherally in muscles ^17^. The DBS-related mEP have been reported by several other investigators to be associated with DBS-induced motor side effects ^24,25^. Activation of the STN-GP pathway has recently been proposed to be reflected in the DBS local evoked potentials (DLEP) ^26^. The DLEP signal, also known as evoked resonant neural activity (ERNA), is recorded from inactive DBS lead contacts around the stimulating contact in the STN and is related to the therapeutic efficacy of DBS ^27–30^.

The goal of this study was to experimentally quantify and compare the activation of DBS-relevant pathways (HDP, CSBT, and STN-GP) during cathodic and anodic DBS in patients with PD (Figure 1). We hypothesized that anodic stimulation is particularly inefficient (achieving less activation with the same amount of current) in activating the side effect-inducing CSBT compared to cathodic stimulation, but this inefficiency is less pronounced when activating the therapeutic HDP and STN-GP pathways. We compare our experimental findings with computational modeling predictions to further elucidate biophysical mechanisms of electrical stimulation in human brain, and with prior clinical observations to shed light on which subthalamic pathways may be contributing to DBS therapeutic benefits.

**Figure 1.**
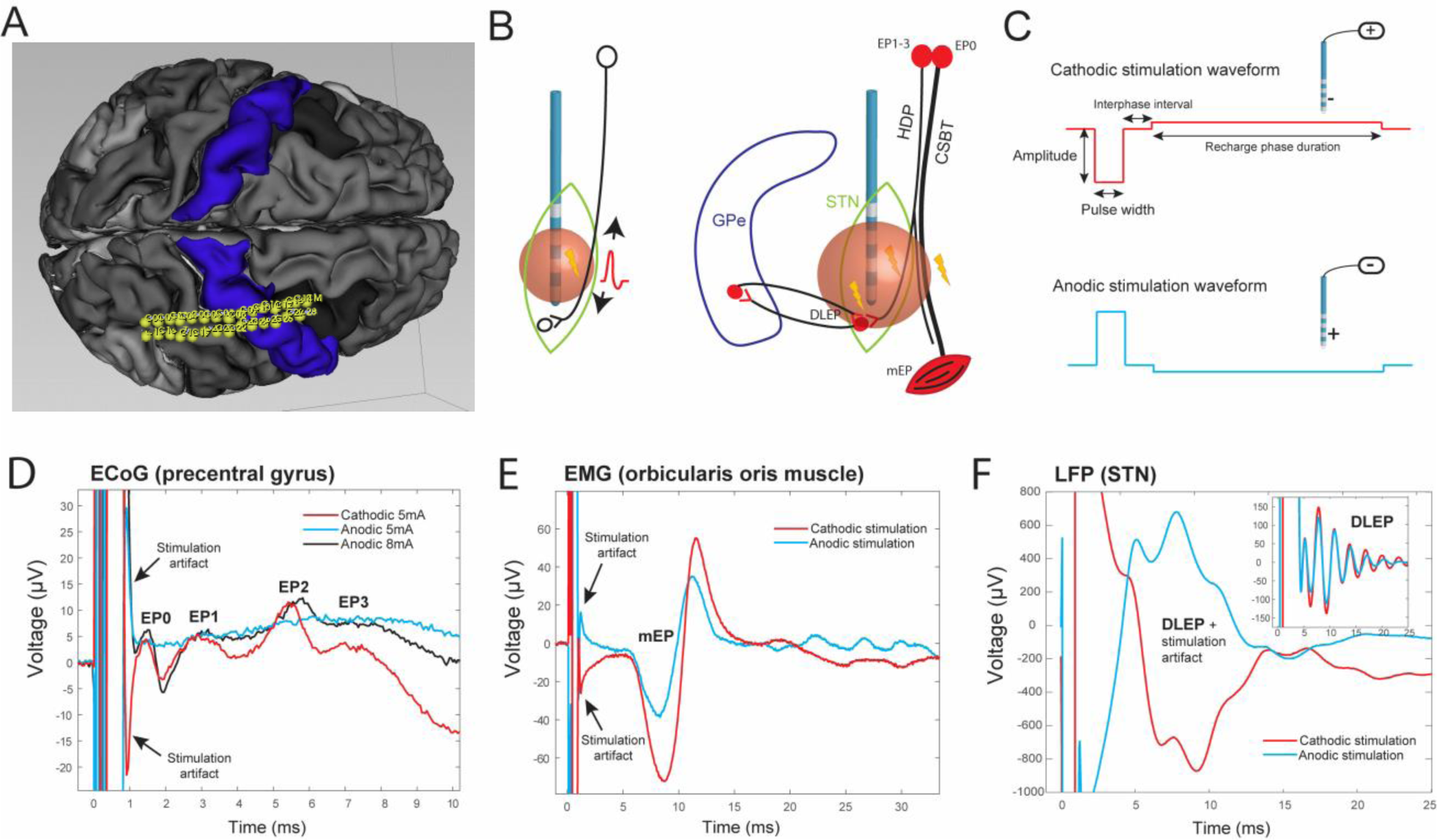
Experimental setup. A) Electrocorticography (ECoG) strip, co-registered with surface-extracted preoperative MRI, was placed over the arm area of the primary motor cortex B) Left: activation of an axon during extracellular stimulation results in antidromic and orthodromic action potential propagation. Right: A schematic of clinically relevant pathways in the STN region and evoked potentials corresponding to their activation. C) Asymmetric biphasic waveform for monopolar cathodic and anodic stimulation (see text for details). D-F: Examples of recorded evoked potentials during cathodic (red) and paired anodic (blue) stimulation for cortical evoked potentials (cEP) (D), motor evoked potential (mEP) (E), and raw and filtered (inset box) DBS local evoked potential (DLEP) (F). Each trace was obtained by averaging signal in response to ∼120 DBS pulses. Stimulation artifact reverses direction while neural or muscle response is consistent for cathodic and anodic stimulation. STN= subthalamic nucleus; GPe = globus pallidus externa; HDP = hyperdirect pathway; CSBT = corticospinal/bulbar tract; EMG = electromyography; LFP = local evoked potential.

## Materials and methods

### Patient selection

Patients with idiopathic Parkinson’s disease scheduled to undergo awake STN DBS surgery at a large academic center were recruited for the study (Table 1). Informed consent was obtained before surgery under the protocol approved by the Institutional Review Board. All patients were informed that a temporary subdural electrocorticography (ECoG) recording electrode would be placed strictly for research purposes during surgery.

**Table 1.** Patient demographics and experimental setup.

| Patient code | Age/Sex | Stimulated hemisphere | ECoG laterality at M1 (mm) | Number of M1/total ECoG channels | Number of face/limb EMG channels | Number of cathodic DBS settings | Number of anodic DBS settings | DBS lead bottom contact MCP coordinates (mm) |  |  | DBS lead model |
| --- | --- | --- | --- | --- | --- | --- | --- | --- | --- | --- | --- |
| P1 | 5X/M | R | 28.0 | 9 / 26 | 3 / 3 | 39 | 5 | 10.7 | -2.5 | -6.6 | BSC 2202 |
| P2 | 6X/M | L | 26.5 | 10 / 26 | 1 / 3 | 34 | 6 | -12.7 | -2.3 | -4.2 | ABT 6172 |
| P3 | 6X/M | R | 23.9 | 11 / 26 | 0 / 4 | 31 | 19 | 12.4 | -2.6 | -2.9 | ABT 6172 |
| P4 | 4X/F | R | 29.0 | 9 / 26 | 2 / 3 | 19 | 4 | 9.6 | -3.3 | -7.9 | BSC 2202 |
| P5 | 6X/M | R | 20.0 | 2 / 5 | 2 / 3 | 40 | 6 | 9.2 | -3.4 | -5.9 | BSC 2202 |
| P6 | 5X/F | R | 24.1 | 1 / 5 | 2 / 3 | 32 | 6 | 11.4 | 0.3 | -4.4 | BSC 2202 |
| P7 | 6X/M | R | 27.6 | 10 / 26 | 3 / 2 | 8 | 7 | 8.1 | -4.4 | -5.7 | BSC 2202 |
| P8 | 4X/M | R | 22.7 | 3 / 5 | 0 / 0 | 4 | 4 | 12.1 | -2 | -6.5 | BSC 2202 |
| P9 | 6X/M | R | 20.9 | 2 / 5 | 0 / 3 | 4 | 4 | 12.5 | -2.9 | -4.7 | MDT B33005 |
| P10 | 4X/M | L | 32.5 | 6 / 26 | 1 / 4 | 24 | 24 | -12.5 | -2.6 | -3.9 | BSC 2022 |
| P11 | 6X/M | R | 32.3 | 7 / 26 | 3 / 4 | 25 | 22 | 9.4 | -3.6 | -6.6 | BSC 2022 |
| P12 | 6X/F | R | 36.6 | 9 / 26 | 4 / 4 | 62 | 60 | 9.0 | -2.5 | -4.4 | BSC 2022 |
| P13 | 5X/M | R | 29.9 | 8 / 26 | 4 / 4 | 41 | 43 | 11.3 | -2.8 | -5.5 | BSC 2022 |
| P14 | 4X/F | R | 25.5 | 7 / 26 | 0 / 4 | 52 | 52 | 8.5 | -4.0 | -6.2 | BSC 2022 |
| P15 | 7X/M | L | NA | NA | NA | 40 | 40 | -11.4 | -3.4 | -7.0 | BSC 2022 |

M1 = primary motor cortex; MCP = mid-commissural point; ABT= Abbott; BSC = Boston Scientific; MDT= Medtronic.

### Research-related surgical methods

Segmented DBS electrodes (Medtronic, Abbott or Boston Scientific models) were implanted using microelectrode guidance and standard clinical procedures ^31^. To record cortical evoked potentials (cEP), an ECoG strip (28 or 6 contacts; Ad-Tech) was placed subdurally through the same burr hole used for DBS implantation ^17,32^. The 28-contact strip had two rows of fourteen 2-mm-diameter platinum contacts separated by 4 mm. The 6-contact strip had one row of 4-mm-diameter platinum contacts separated by 10 mm. The intended target location for the center of the strip was the arm area of M1, ∼3 cm from the midline and slightly medial to the hand knob. The motor evoked potentials (mEP) were recorded using surface EMG electrodes placed on the contralateral arm (biceps, flexor carpi radialis, extensor carpi radialis, and first dorsal interosseous), and bilateral face (nasalis, orbicularis oris). The DBS electrode was used to deliver stimulation and to record DBS local evoked potential (DLEP) activity from non-stimulating contacts. Recordings were performed at least 12 hours after stopping all anti-parkinsonian medications and at least 30 minutes after stopping propofol sedation.

### Stimulation

Electrical stimulation was delivered through the DBS electrode contacts while patients were at rest using the NeuroOmega electrophysiology system (Alpha Omega Engineering). The number of stimulation settings tested varied between patients depending on the duration of time available intraoperatively (average 50, range 8–122). The stimulation settings varied in active contact, amplitude, pulse width, and stimulus polarity, and were delivered in a pseudorandomized order (*randperm* function in MATLAB). All stimulation settings were monopolar and 2×3 inch surface electrode on the shoulder ipsilateral to the stimulated brain hemisphere was used as the return.

Stimulation frequency was 10 Hz (except 9Hz for P08) and was applied for 12 seconds (13.3 seconds for P08) with 3 seconds pause between the settings. In 3 patients (P12, P13, and P15), stimulation settings below 1 mA were applied for 3 seconds with 1 second pause specifically to elicit DLEP (DLEP measurements required fewer pulses as the signal-to-noise ratio was typically high). The stimulation waveform was asymmetric biphasic with 70 us inter-phase duration with the first, large-amplitude phase that defined stimulation polarity (cathodic or anodic), and similar to a waveform delivered by clinical stimulators ^33^. The most common stimulation pulse width was 60 us so the second phase was typically 8 times longer with 1/8 of the amplitude of the first phase to maintain charge balance between phases. Because of the 500 us NeuroOmega limit for each phase duration, stimulation settings with pulse widths longer than 60 us had their second phase duration limited to 480 us and second phase amplitude adjusted to maintain charge balance. Consequently, for some stimulation settings, the amplitude of the second phase could be large and might contribute to neural activation. We, therefore, excluded from analysis any stimulation settings where second phase amplitude was over the EP activation threshold (400 us anodic settings for P11, P12, and P13 and 400 us cathodic settings for P13). Because long pulse width settings were necessary to construct the strength duration curves described below, in two patients (P14, P15) we utilized the analog stimulation feature of NeuroOmega where a waveform of arbitrary shape (including phase duration) can be defined. For these stimulation settings, the duration of the second phase was variable depending on the first phase pulse width, and up to 10ms.

### Signal Acquisition

ECoG, local field potentials (LFP) and EMG signals were recorded simultaneously with the NeuroOmega. Signals were amplified and acquired at 22 kHz sampling rate with a built-in hardware bandpass filtering between 0.075 and 3500 Hz. The contra- and ipsilateral ear lobes were used as the ECoG reference and ground, respectively. The EMG signals were recorded from pair-referenced electrodes and a common ground was on the contralateral knee. The LFP was recorded from all non-stimulating DBS contacts. An ipsilateral scalp needle was used as a recording reference while corresponding contralateral scalp needle served as the ground for LFP.

### Electrode localization

The ECoG electrode location was determined using intraoperative CT registered to the preoperative MRI with cortical surface extracted in the FreeSurfer or Fastsurfer software v7, and visualized in Slicer 5.2.2. EcoG contacts over the precentral gyrus representing the primary motor cortex (M1) were selected for further analysis. On average, 8 (range 1-11) ECoG bipolar contact-pairs were used per patient (Table 1). For localization of the DBS lead, we used the CranialSuite software 6.3.1 (Neurotargeting). Briefly, postoperative CT scans were linearly coregistered to preoperative T1-weighted MRI scans. The electrodes position was automatically detected and manually verified. The coordinates of the most ventral contact are reported relative to the mid-commissural point (MCP) (Table 1).

### Signal Processing

#### ECoG (cEP) analysis

Data analysis was performed using custom scripts in MATLAB vR2021b (MathWorks). Raw ECoG potentials were re-referenced in a bipolar montage using adjacent contacts, aligned by stimulus start times, and averaged to generate cEP (approximately 120 pulses for each DBS setting). The averaged signal was baseline-shifted to zero using 1 ms of data before stimulation pulse, and a smoothing function (5-point window moving average) was additionally applied (Figure 1-D). The cEPs were classified based on the temporal order of appearance and according to the peak latencies with respect to stimulation onset as previously defined ^17^ : EP0 (< 2 ms), EP1 (2-3.5 ms), EP2 (3.5-7 ms), and EP3 (7-10 ms). The EP amplitude was defined as the voltage difference between EP peak and its preceding trough or baseline. The presence or absence of peaks and their amplitudes depended on stimulation settings while peak latencies were relatively consistent especially within the same patient ^17^.

Given the large number of stimulation settings and recording channels, we developed a novel algorithm for semi-automated detection of cEP peak latencies and amplitudes. The algorithm first defined a template signal for each channel by averaging the normalized responses evoked by the highest stimulation current. Then, using the prominence and width concepts in the *findpeak* function in MATLAB, the baseline, peaks, and troughs of the template were determined in a recursive and supervised manner. The algorithm then searched for the baseline and the extremum points of the evoked potential within the width of the template. All algorithm selections were visually confirmed. In cases where baseline could not be clearly identified due to the large stimulation artifact, EP1 was not defined (6% of the recordings).

The accuracy of the algorithm was confirmed by comparing with previously reported manually extracted EP1 amplitudes ^34^. The average correlation coefficient between the algorithm and manual assessment was 0.92±0.04, with 5.1±2.0% of algorithm detections requiring user revision. Given the large amount of data requiring visual inspection even with the semi-automated approach, we identified one best representative channel to report for EP2 and EP3 analysis (a channel with minimal noise and the largest cEP amplitudes for both anodic and cathodic stimulation, with anodic response prioritized if necessary). The EP0 was extracted manually since automated detection could not be implemented given its variable proximity to the stimulation artifact (one best channel was selected for this analysis).

#### EMG (mEP) analysis

Similar to the cEP analysis, EMG responses were aligned by stimulus start times and averaged to generate mEP for each recorded muscle in response to each stimulation setting (Figure 1-E). The presence of mEP was determined by visual inspection as a clear deflection from the baseline at expected latency (∼10 ms for facial muscles, 15-30 ms for arm muscles), and peaks and troughs were manually annotated. The mEP amplitude was defined as the largest peak-to-trough distance (mEP could be multiphasic with multiple peaks and troughs). For each patient, the best muscle was defined as the one with the most mEP responses. In most patients this also corresponded to the muscle with the largest mEP amplitudes, and for all but three patients, it was also the channel that correlated best with EP0.

#### DLEP analysis

The LFP signals were recorded in a monopolar montage from all non-stimulating contacts on the DBS electrode. As with other EP types, stimulation times were aligned and LFP signal averaged. Given the close proximity of recording contacts to the stimulating contact, a large stimulation artifact was present. To remove the stimulation artifact and extract DLEP signal from all recording contacts, we developed a novel processing pipeline, based on fitting the stimulation artifact with a family of damped sinusoids and a post-smoothing step with a Savitzky–Golay filter. With this approach, the stimulation artifact was modeled as the compounded step response (the stimulation waveform) of the transfer function of a second-order system^35^ (Figure 1-E). The DLEP amplitude was defined as the root mean square (RMS) of the artifact-free signal in the 4-20 ms post-stimulation recording window. For each stimulation setting, we calculated the average DLEP amplitude from all contacts/segments immediately adjacent (dorsal and ventral) to the stimulating contact. DLEP latency was defined as the latency of the first peak after applying the filtering.

### Pathway activation analyses

#### Recruitment curves

The recruitment curve (RC) graphically represents the relationship between the stimulation current intensity and the corresponding neuronal response (EP amplitude) while holding all other stimulation parameters constant (stimulus polarity, active contact, pulse width). The RCs were constructed for each EP type, both for individual patients and on a group level. To allow comparison of EP amplitudes on a group level, EP amplitudes were normalized from 0 to 1 (the largest amplitude during 60us pulse width stimulation was defined as 1 since 60us settings were used in all patients). For mEP, the normalized values for each muscle were also averaged to obtain one mEP amplitude for each stimulation setting. For group curves, we used only stimulation settings with the bottom active contact and 60 us pulse width (P09 excluded since 60us stimulation was not applied). At least two patients had to contribute data for a specific stimulation amplitude to be included in the group analysis.

#### Strength-duration curves

Given that different neuronal elements (e.g. fibers of passage, local neurons) have different strength–duration (SD) characteristics, we constructed SD curves for all EP with direct neuronal origin (excluding mEP) in 5 patients (P11-P15). In each patient, we tested settings sweeping the pulse width from 30 us to 400 us. For each pulse width, we applied an ascending range of current amplitudes (5 on average) for both cathodic and anodic polarity. To find the activation current threshold for each pulse width, we constructed a recruitment curve for each EP measure. The stimulation current at the first inflection (knee) point of the recruitment curve was selected as the activation threshold. If the knee point was not present, we chose one EP value for both cathodic and anodic recruitment curves and defined corresponding stimulation currents as the activation current thresholds. The SD curves were constructed by plotting pulse widths (duration) on the x-axis and activation thresholds (strength) on the y-axis. From each SD curve, we then calculated the chronaxie metric, a time constant closely related to the time constant of the firing neuronal element ^36^. We used Weiss’s linear relationship ^37,38^ between threshold charge (pulse-width * stimulation-current) and pulse-width (duration) to estimate the chronaxies. Specifically, chronaxie time was defined as the intersection with the pulse width (x-axis), and calculated by dividing the y-axis intercept value by the slope ^37,39,40^.

#### Excitability metric

To compare pathway excitability during cathodic or anodic stimulation, we calculated area under the curve (AUC) of the averaged normalized group and individual patient recruitment curves. Different metrics have been used to characterize the stimulation response in prior studies including the AUC, slope of the original recruitment curve, or slope of a fitted sigmoid curve ^41–44^. We chose AUC as our RC characterization metric since we did not have enough data points for each patient to reliably extract a slope or fit a sigmoid function (we could not determine if RC curves reached the saturation limit, which is necessary to calculate the slope or fit a sigmoid curve). To calculate the AUC, we used trapezoidal integration method in which the total area is first divided into smaller trapezoids between each successive data point and then summed up. For both cathodic and anodic RC, the AUC was calculated from 0 mA to the highest stimulation amplitude available for both polarities (typically 6 mA).

### Statistics

We compared EP amplitudes and latencies between paired stimulation settings (same stimulation parameters except waveform polarity). To avoid collinearity between recorded channels, we selected the best representative channel for each EP and calculated the difference between anodic and cathodic amplitudes. Then, we fit Gaussian generalized estimating equation (GEE) models to the difference’s values with exchangeable correlation structure and jack-knife variance estimates^45^.

This robust approach accounts for possible correlations between observations at different settings in the same patient. Then the intercept in these models tests the null hypothesis that there is no difference between cathodic and anodic settings. We repeated this analysis for EP latencies. We also examined EP1 in all M1 channels for each patient, in which we used Wilcoxon signed-rank tests for each patient (treating channel and setting as the unit of replication) and a Bonferroni correction to account for the number of tests in this sub-analysis (i.e., number of patients). To compare the area under the curve (AUC) of cathodic and anodic recruitment curves in individual patients, we again used non-parametric Wilcoxon signed-rank test. Similarly, we used the Wilcoxon rank test to compare cathodic and anodic current threshold for HDP activation (associated with EP1). A p-value <0.05 was considered statistically significant.

## Results

We recorded subdural electrocorticography (ECoG), local field potentials (LFP) and electromyography (EMG) signals in 15 patients with Parkinson’s disease in response to low-frequency cathodic or anodic monopolar DBS in the STN (Table 1; Figure 1). The signals were epoched and averaged to reveal evoked potentials associated with antidromic or orthodromic activation of neural pathways in the STN region. The very short latency (< 2ms) cortical EP (EP0) and peripherally recorded mEP were a metric for activation of CSBT; short-latency (2-10ms) cortical EP (EP1, EP2, and EP3) for activation of HDP; and DLEP for activation of STN-GP pathway. On average we tested 30 (range 4-62) cathodic and 20 (4-60) anodic simulation settings for each patient depending on the available intraoperative experimental time. We varied the active contact, stimulation amplitudes and stimulation pulse width, while frequency was set to 10 Hz to allow EPs to be recorded between the pulses. We tested a total of 157 paired stimulation settings (average 10 per patient; range 4-32) meaning all stimulation parameters were the same except for the waveform polarity, allowing direct comparison of EP responses for cathodic and anodic stimulation.

### Anodic stimulation is less efficient than cathodic stimulation

For EP1, EP2, EP3, EP0, and mEP the response amplitude evoked by cathodic stimulation was significantly larger compared to the amplitude evoked by its paired anodic stimulation (Figure 2; Table S1). The DLEP amplitude did not significantly differ between cathodic and anodic stimulation (p-value=0.63). The same pattern of higher amplitudes with cathodic compared to anodic stimulation held true when we inspected EP1 responses in all M1 channels (Figure S2) and analyzed M1 channels in patients individually (Table S2). The normalized mEP over all channels (and not only the best channel) was also consistently larger for cathodic stimulation (Figure S2).

**Figure 2.**
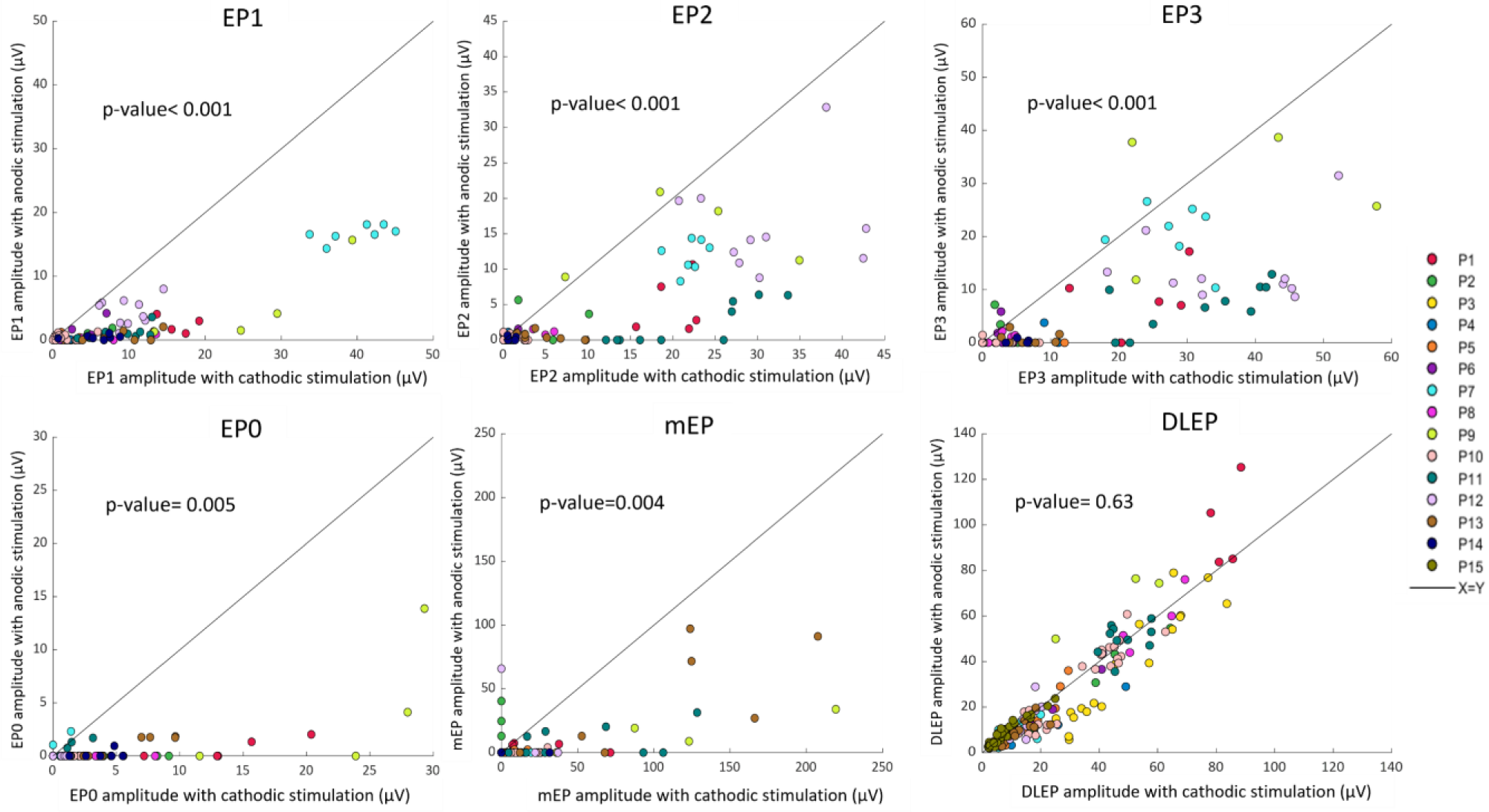
Comparison of evoked potential amplitudes in response to cathodic and anodic stimulation (paired stimulation settings). Each point represents EP response for one stimulation setting pair, colored by patient. Response from one ‘best’ recording channel (largest response) is shown for clarity. The p-values indicates significant differences in all pairwise comparisons with most responses lying below the unity line indicating larger EP amplitudes with cathodic stimulation, except for DLEP.

To investigate whether both anodic and cathodic EP responses had the same morphology, we additionally compared the latencies of cathodic EP peaks with their paired anodic latencies (Figure S1 and Table S1). The latencies of cathodic and anodic peaks followed the same predefined millisecond ranges (Figure 1). However, on average, we observed a small delay (100-500 us) for anodic stimulation in EP2, EP3, and EP0 (p-value<0.01), while delays for EP1, mEP, and DLEP were not significant.

### Relative excitability comparing cathodic and anodic stimulation is pathway-specific

To compare relative excitability of different pathways during cathodic or anodic DBS, we generated group recruitment curves using normalized EP amplitudes (Figure 3). The area under the curve (AUC) quantifies the degree of activation while the ratio between cathodic and anodic AUC provides a measure of relative excitability (higher ratio indicates that cathodic stimulation activates a pathway more than anodic). The highest AUC ratio was observed for EP0 and mEP at 11.8 and 12.2, respectively. Mid-range AUC ratios were measured for EP1, EP2, and EP3 at 8.2, 6.6, and 5.3, respectively, while DLEP AUC ratio was the lowest at 1.1.

**Figure 3.**
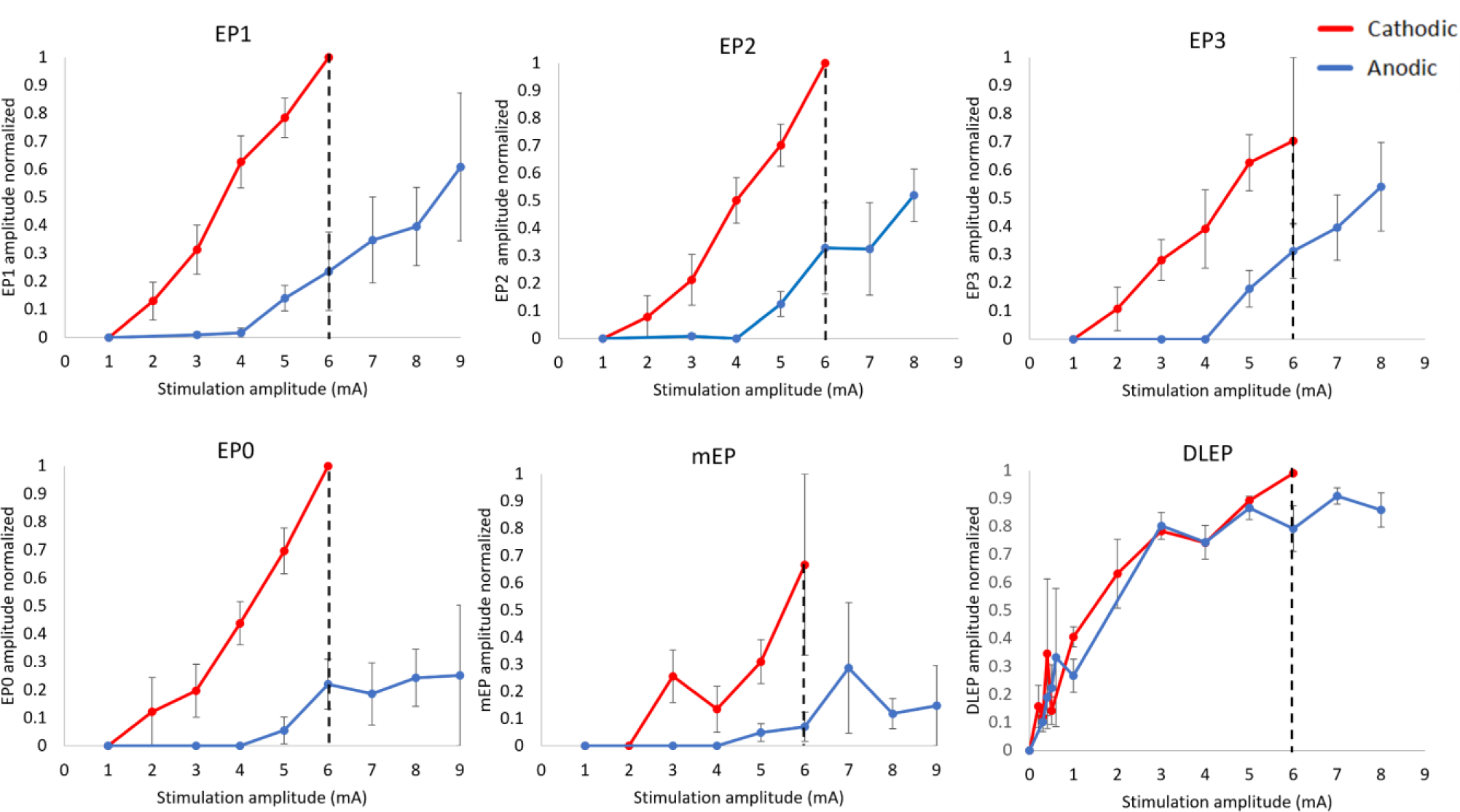
Group recruitment curves for all EP during cathodic (red) or anodic (blue) stimulation. The recruitment curves were generated by averaging normalized EP amplitudes during stimulation at the ventral-most active contact and 60 us pulse width. Error bars indicate standard error. The dotted line at 6 mA indicates the highest common stimulation amplitude up to which the area under the curve (AUC) was calculated. Error bars indicate ± 1 standard error (SE).

We conducted a similar analysis between AUC of cathodic and anodic recruitment curves for individual patients (Figure S3). The AUC values were significantly higher during cathodic compared to anodic stimulation for all EP except DLEP (Table S3). The ratios of average cathodic-to-anodic AUC followed the same trend as for the group recruitment curves with EP0 and mEP the highest (14.8 and 14.6), followed by EP1, EP2, and EP3 (5.5, 4.6, and 3.9) and then DLEP (1.1). This is consistent with the group AUC analysis and the difference between EP0 and EP1 ratios is even more pronounced.

### Experimental excitability metrics are consistent with computational modeling predictions

We compared recruitment curves based on a recent STN DBS computational modeling study with the corresponding curves from our experimental data (Figure 4). For the computational modeling curves, we reformatted data reported in ^13^ and plotted the percentage of activation for CSBT and HDP pathways in response to cathodic and anodic ascending currents (Figure 4-A). For comparison, we plotted on the same graph the recruitment curves reported in the previous section for EP0 and EP1, as respective evoked potentials associated with CSBT and HDP activation (Figure 4-B). In general, computational predictions reached full (100%) pathway activation at lower current amplitudes compared to our EP measures which did not appear to reach saturation. To allow a comparison between modeling and experimental results, we chose 1 mA cutoff to calculate AUC for the model since this corresponds to an approximate 50% activation point. The cathodic-to-anodic AUC ratios for the CSBT and HDP were 8.05 and 2.22, respectively, for the model compared to 11.8 and 8.2 for experimental EP0 and EP1. While the ratio values are not the same, the consistent finding between model predictions and experimental results is that the CSBT excitability metric is higher compared to the HDP excitability metric.

**Figure 4.**
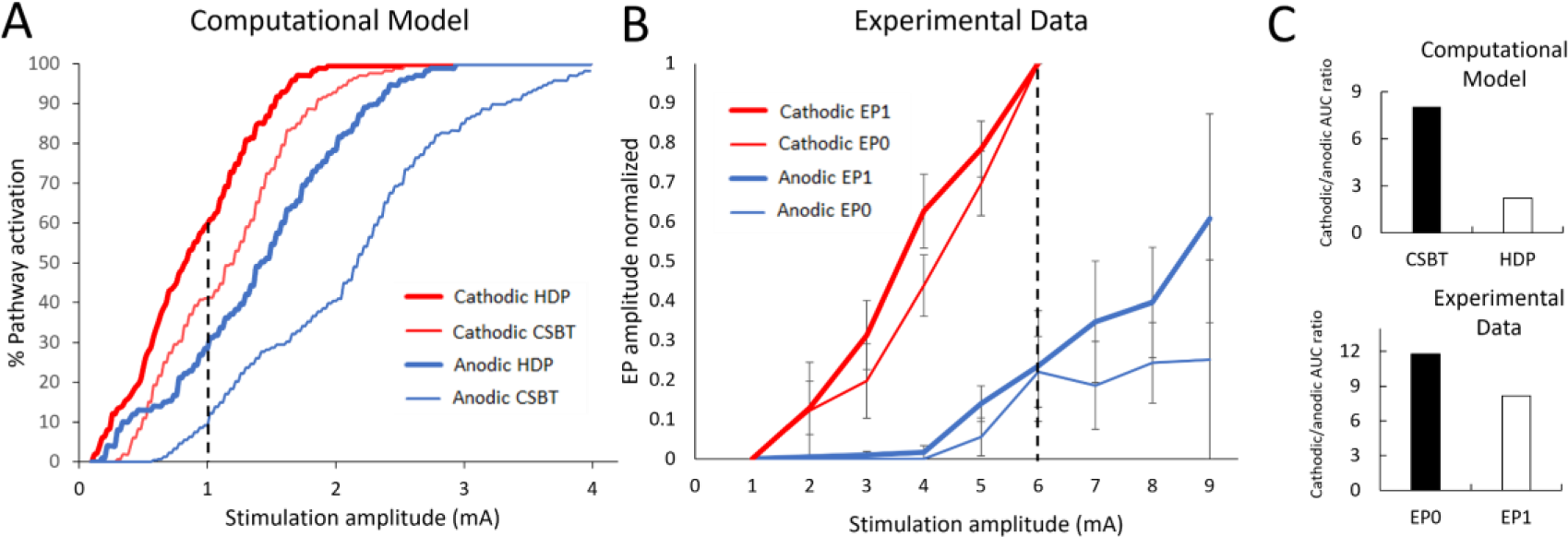
Comparison between computational modeling and experimental recruitment curves for cathodic (red) and anodic (blue) DBS. A) Recruitment curves for percent activation of HDP (thick lines) and CSBT (thin lines) pathways from a computational modeling study (adapted from (Bingham & McIntyre, 2022)) B) Recruitment curves for experimental EP1 (thick lines) and EP0 (thin lines) amplitudes. The dashed line illustrates maximum current for AUC calculation. C) Comparison between cathodic/anodic AUC ratio for modeling (top) and experimental data (bottom).

### Anodic stimulation activates the same neuronal elements as cathodic stimulation

In order to investigate which neuronal elements were first activated during stimulation, for a subset of patients (N=4 for cEP; N=2 for DLEP), we constructed strength-duration curves for all evoked potentials with direct neuronal origin (all except mEP) and calculated corresponding chronaxie times (Figure 5). This required varying pulse width from 30 us to 400 us and testing on average 5 stimulation amplitudes to find an approximate EP activation threshold for each pulse width, and for each polarity. For all evoked potentials, the calculated chronaxie values for both cathodic and anodic stimulations were comparable and within the same range of 80-140 us for cEP and ∼190 us for DLEP (Figure 5). Formal statistical testing was not feasible due to the low number of data points, but nonetheless chronaxie values in this range are consistent with activation of myelinated fibers ^4,46^. These chronaxie values are also consistent with biophysical models of STN neuron activation with DBS ^47^.

**Figure 5.**
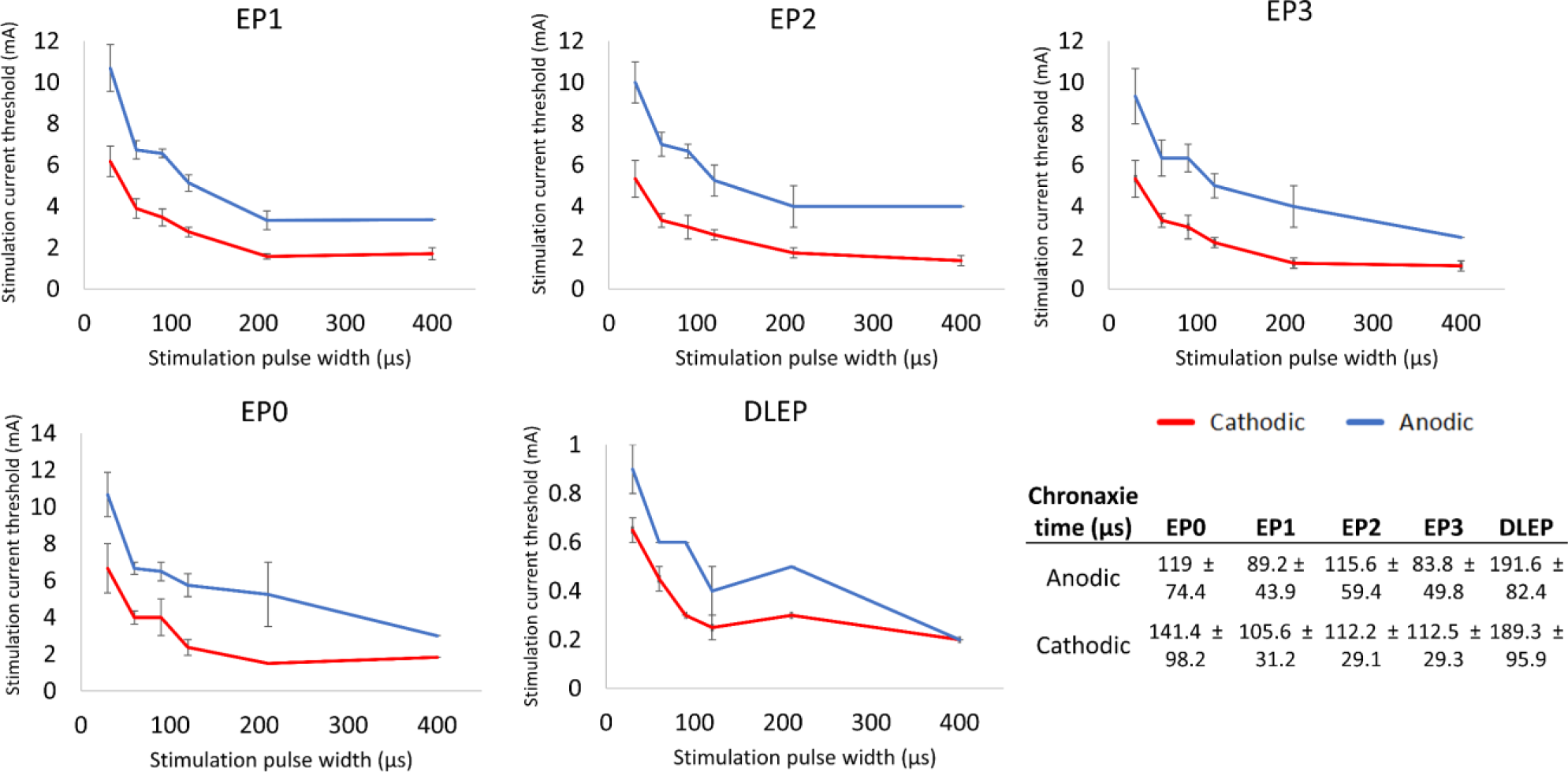
Strength-Duration curves for evoked potentials with neuronal origin. The average chronaxie times extracted from individual patient’s SD curves are reported in the table. N=4 for cEP; N=2 for DLEP (DLEP 400us point is based on 1 patient only). Error bars indicate ± 1 SE.

### Anodic stimulation requires higher current amplitude for similar degree of activation as cathodic stimulation

Given that anodic stimulation was consistently less efficient in activating HDP and CSBT pathways, we asked what current amplitude is needed for anodic stimulation to achieve the same activation as cathodic stimulation. Using individual patient recruitment curves (Figure S3), we extracted the current stimulation amplitude needed to evoke EP1 of 1uV as a threshold for HDP activation. We used data from 7 patients who had sufficiently detailed recruitment curves (3 or more data points) for both polarities and current thresholds were averaged across all M1 channels. On average, the cathodic current threshold for HDP activation was 2.46 ± 0.8 mA compared to 5.26 ± 1.4 mA for anodic stimulation (p-value= 0.031). The average anodic/cathodic current ratio was 2.2 ± 0.44, indicating that more than twice the anodic current was needed to activate the HDP (Table S4).

## Discussion

We compared evoked potentials attributed to activation of clinically-relevant pathways in the subthalamic region in response to anodic and cathodic DBS. We studied intracranial and peripheral (muscle) responses in 15 patients with PD. By focusing on stimulus polarity, our goal was to elucidate biophysical mechanisms that underlie electrical stimulation in the human brain. Our key observations are: 1) subcortical cathodic and anodic stimulation evoke similar responses in the cortex, subcortically and peripherally, but anodic responses are generally smaller for the same current amplitude; 2) relative efficiency of anodic stimulation compared to cathodic varies by EP measure meaning that anodic stimulation is the least efficient in activating CSBT (EP0/mEP), moderately efficient in activating the HDP (EP1/EP2/EP3), and as efficient as cathodic in activating STN-GP pathway (DLEP); 3) cathodic and anodic stimulation have comparable chronaxie times for all EP measures; 4) relative excitation efficiency for experimental EP0 and EP1 markers is consistent with computational studies modeling activation of CSBT and HDP pathways, respectively; 5) anodic stimulation requires approximately twice as much current as cathodic to achieve the same degree of EP1 (HDP) activation.

### Cathodic and anodic stimulation activate the same neural pathways

EP measures that we utilized in this study have previously been attributed to activation of specific pathways in the subthalamic region ^16–26,28,29,48–50^. In this study, we demonstrate that the morphology of evoked responses is similar during cathodic and anodic stimulation suggesting that the same pathways are activated regardless of stimulus polarity. Specifically, we observed the same number, shape and general timing of peaks in the cortex, subcortically and peripherally, however with anodic stimulation the peaks were smaller and, in the cortex, sometimes delayed by up to 0.5 milliseconds.

From physiological studies in the peripheral and central nervous system, it is known that stimulating an axon fiber passing by a monopolar electrode requires less cathodic current than anodic ^4,46,51^. This is because electric field for cathodic stimulation generates a large depolarization near the electrode with small hyperpolarizations flanking it. In cable theory, this is expressed mathematically as the ‘activating function’ stating that membrane polarization and activation of a straight axon (a cable) depends on the second order spatial derivative of the extracellular electric potential ^52–54^. For anodic stimulation, the electric potential along a passing axon is reversed so that there is a large hyperpolarization near the electrode and small depolarizations at the flanking regions (Figure 6 C). As a result, a larger anodic stimulus is needed to achieve suprathreshold activation at the depolarized flanking regions. During cathodic stimulation, an action potential can propagate through hyperpolarized regions if hyperpolarization is small enough, but a large hyperpolarization can extinguish it, known as the ‘anodal surround’ ^4^ (Figure 6D). When monopolar electrode is near a cell body, rather than along an axon, such as during cortical stimulation, the lowest threshold is usually anodal because, due to neuron orientation with respect to the electrode, the current hyperpolarizes the dendrites and cell body and depolarizes the axon hillock (Figure 6A) ^4,55^.

**Figure 6.**
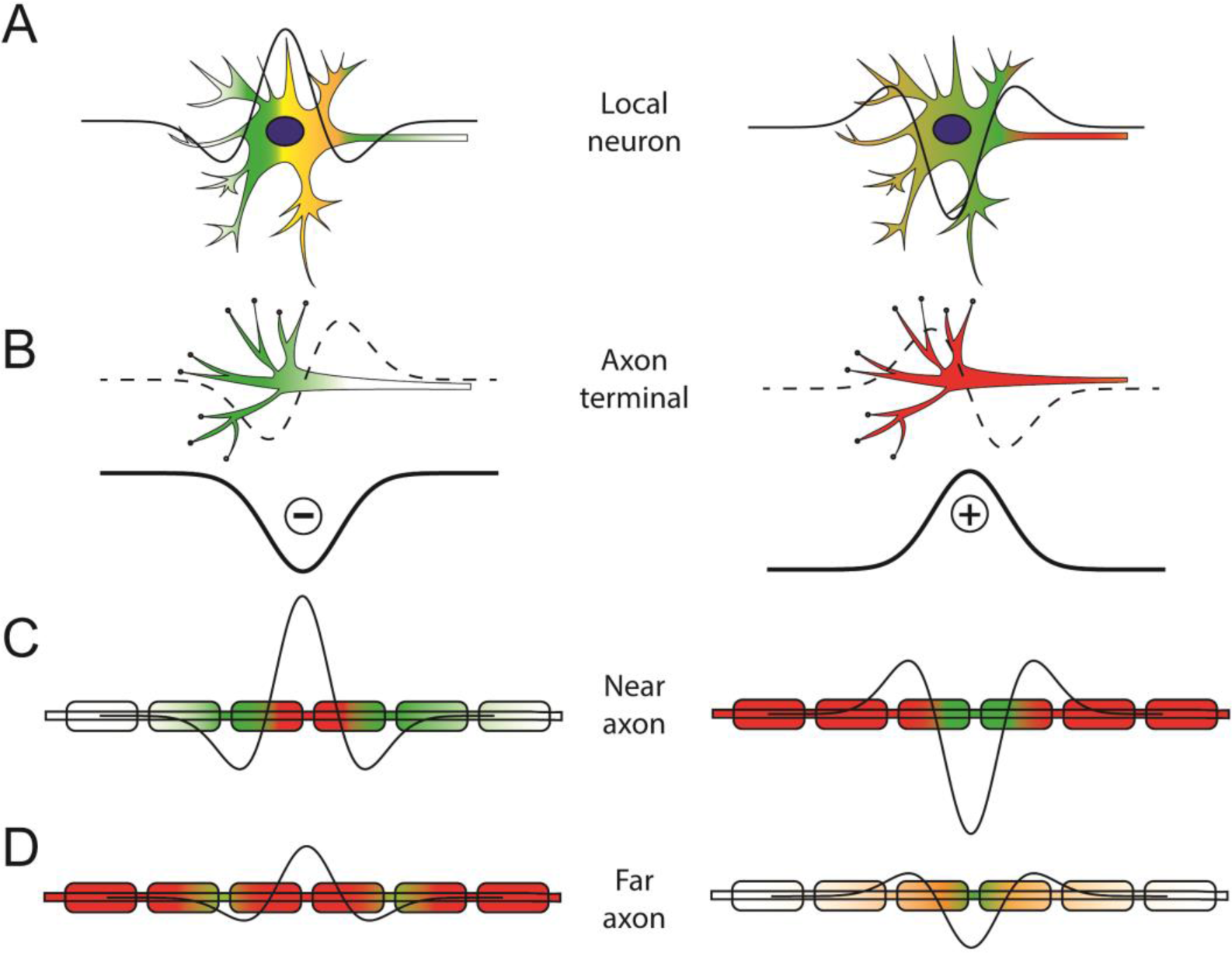
Comparison between activation of different neuronal compartments in response to cathodic (left) and anodic (right) stimulation. Depolarization is depicted in warm colors while hyperpolarization is in green. Thick line shows extracellular potential around monopolar cathode (left) or anode (right) decaying with distance from the electrode. Action potential initiates in the region with over-the-threshold depolarization (red). When cell body of a local neuron is the closest compartment to the electrode (A), activation occurs at the axon hillock as sufficient direct depolarization of cell body is difficult to achieve with extracellular stimulation. Activation of axon terminals (B) is regulated by the first spatial derivative of the electric potential which is symmetric for cathodic and anodic stimuli (dashed line). Activation of passing axons (including the axon hillock) is driven by the second spatial derivative of the electric potential (thin line; a.k.a. the activating function). Axons closest to the electrode (C) may not be activated by cathodic stimulation due to strong hyperpolarization blocking the propagation of action potential, while those farther away (D) are most excitable by cathodic stimulus. Note that activation of neural elements by extracellular electric field does not depend solely on the value of its first or second spatial derivative but rather it is driven by the precise timing and spatial distribution of induced currents exiting and entering neural compartments and leading to their depolarization or hyperpolarization. This is a dynamic process that cannot be easily captured in a schematic but the purpose here is to provide a visual guide highlighting relevant biophysical concepts.

### Cathodic and anodic stimulation activate the same neural elements

To identify the primary neural elements related to activation of different pathways, we constructed strength-duration curves for all EP measures and extracted chronaxie time constants. Smaller chronaxie values indicate more excitable neural elements ^56^. Previous studies have estimated the chronaxie values for large, myelinated axons to be ∼30-200 ms, for small axons ∼200-700 ms, and for cell bodies and dendrites ∼1-10 ms ^4,37,40^. In our study, the chronaxies for all EPs were comparable for cathodic and anodic stimulation and fell within the range associated with axonal activation. We found that the average chronaxie values for DLEP (∼200 us) were longer, which might be an indicator of a mixture of other neuronal elements being activated for that pathway, such as terminal synapses or axon hillock ^4^.

### Efficiency of cathodic compared to anodic stimulation is pathway-specific

One key finding of this study is that the relative activation efficiency (amount of activation per unit of current) of cathodic compared to anodic stimulation varies by the pathway. This has important implications regarding how precisely pathway activation initiates when extracellular stimulation is applied subcortically. Similar chronaxies suggest that activation occurs in the same axonal element regardless of polarity, but differences in relative efficiencies suggest that different axonal elements may be involved for different pathways. We propose that differences in relative activation efficiency arise from different pathway distances from the stimulating electrode and variations in which neural elements are exposed to the electric field given the specific local anatomy of this brain region.

The CSBT is composed of large, myelinated fibers that pass parallel to the stimulating electrode usually at some distance from the electrode, and the goal of clinical STN DBS is to avoid CSBT activation and associated side effects. Since the axons are passing by the electrode, the activation of CSBT is driven by the second spatial derivative of the extracellular potential in which case anodic stimulation is much less efficient ^4^. This was demonstrated in our study for EP0 and mEP which both measure CSBT activation (antidromically for EP0 and orthodromically for mEP). Similar observations were made recently by Campbell et al. (2023) where anodic monopolar STN DBS resulted in much smaller mEP compared to cathodic ^25^.

The HDP is a direct input from the cortex to the STN formed primarily by thin collaterals from corticofugal axons passing through the internal capsule and continuing towards the brainstem, although some may be direct connections ^57–59^. In our study, anodic stimulation was more efficient at activating the HDP compared to CSBT (lower cathodic/anodic activation ratio) (Fig. 3). This suggests that HDP activation involves activation of axonal elements that differ from the distant, large, myelinated passing fibers. Given the anatomical location of the stimulating electrode in the STN, the HDP axons are at varying distances from the electrode as they enter the same target area. Paradoxically, the axons that pass the closest to the electrode are not activated by suprathreshold cathodic stimulation due to anodal surround phenomenon ^4^. That means that anodic stimulation would gain some advantage in this situation over cathodic. Furthermore, the activation of axon terminals is predicted to be driven by the first spatial derivative of the extracellular voltage because of the biophysical properties of the final segment of the multi-compartment cable model ^15,54,60,61^. Consequently, axon terminals are equally likely to be activated by anodic or cathodic stimulation given the symmetric spatial distribution of the electric field (Figure 6-B). As a result, we propose that HDP activation is likely achieved by exciting passing axons at varying distances from the electrode as well as by activating terminal axons synapsing in the target region ^62^.

Anodic stimulation is relatively more efficient at activating HDP compared to CSBT, but overall cathodic stimulation is more generally efficient. Comparison of cathodic and anodic stimulation in rodent hippocampus similarly demonstrated that the two polarities activate different axonal sub-populations in the same pathway ^63^. Different latencies observed for EP2 and EP3 can be explained by HDP being composed from axonal bundles of different sizes as shown in histologic studies ^58^. These structural differences influence axonal excitability and conduction velocity ^64^. Alternatively, EP2 and EP3 could represent recurrent activation of surrounding cortical neurons after the initial antidromic invasion of layer V cells ^65,66^.

The DLEP is thought to result from activation of STN-GPe projections inducing resonant activity between these two nuclei that are reciprocally connected with excitatory (STN-to-GPe) and inhibitory (GPe-to-STN) projections ^26,67^. In contrast to all the other EPs that we studied, the DLEP anodic excitability was very similar to DLEP cathodic. There are two neural elements where anodic activation has an advantage over cathodic: terminal synapses and cell bodies (Figure 6-A&B). The terminal axons present in the STN consist of excitatory cortical projections (HDP) and inhibitory afferents from the GPe which together can generate DLEP activity ^26^. The STN local neurons which project to the pallidum are also directly activated by the stimulating electrode ^68^. Neurons with cell bodies in proximity to the active electrode can be activated with lower thresholds using anodic stimulation due to the depolarization of the axon ^4,54,55,69,70^. As a result, the activation of STN-GPe pathway and DLEP generation is likely due to combined activation of terminal synapses converging in the STN and activation of local STN projection neurons.

### Experimental findings confirm some biophysical DBS model predictions

For several decades computational studies utilizing multi-compartment neuron models have predicted that neural activation thresholds depend on the spatial position and orientation of the neuron relative to the electrode, in addition to the electrical properties such as type and distribution of ion channels, as well as capacitance and conductance of neural compartments ^54,69,71^. These models have been instrumental in helping explain how extrinsic stimulation, such as DBS, engages with the nervous system ^26,33,64,72–74^.

Today, the most advanced computational DBS models utilize representations of individual cortical projection neurons with complex axonal arbors that mimic histological reconstructions to predict activations of HDP and CSBT pathways ^13^. This level of detail is important because activation thresholds depend on neuronal morphology ^15^. Compared to cathodic monopolar stimulation, detailed models predict higher activation thresholds with anodic for both pathways, as well as the longer conduction latencies to the cortex and within the STN ^75^. Furthermore, the model predicts that relative efficiency of anodic stimulation is pathway specific (anodic is relatively efficient at activating HDP but less efficient for CSBT) which is consistent with our experimental data. This phenomenon can be attributed to the greater distance of the CSBT from the stimulating electrode compared to the fibers in the HDP, and likely involvement of terminal axons in HDP activation ^62^.

A recent modeling study by Anderson et al. has suggested that anodic stimulation should be more efficient than cathodic when activating neurons orthogonal to the electrode (leaving or approaching the electrode so that terminal synapse or cell body is closest to the electrode) compared to passing axons ^14^. The authors have predicted that because HDP approaches the DBS electrode orthogonally, the HDP activation will be more efficient during anodic compared to cathodic stimulation. Our experimental data does not support more efficient activation of the HDP, but rather, the STN-GP pathway (DLEP marker) was similarly excitable by anodic and cathodic stimulation. So, while the modeling principles are valid, we suggest that they apply to a different pathway.

### Differences in pathway excitability shed light on clinical DBS mechanisms

There are many pathways coursing through the STN region, and it is still unclear which pathways are responsible for therapeutic benefit in STN DBS ^76^. Specifically, there has been an ongoing debate regarding the relative contributions of the HDP and subthalamo-pallidal pathway (STN projections to GPi and GPe are part of the indirect pathway) ^77^. It is widely accepted that CSBT is the pathway causing motor side effects ^11,24^. During clinical DBS programming, two thresholds are typically determined by gradually increasing stimulation amplitude. The therapeutic threshold represents stimulation amplitude at which therapeutic benefit emerges (e.g. tremor reduction) while side effect threshold is the stimulation amplitude where adverse effects are observed (e.g. muscle contractions). The therapeutic window is the difference between the therapeutic and side effect thresholds (Figure 7). Chronic stimulation is typically applied at electrode contact with the largest therapeutic window so it is desirable to devise stimulation strategies that can widen the therapeutic window.

**Figure 7.**
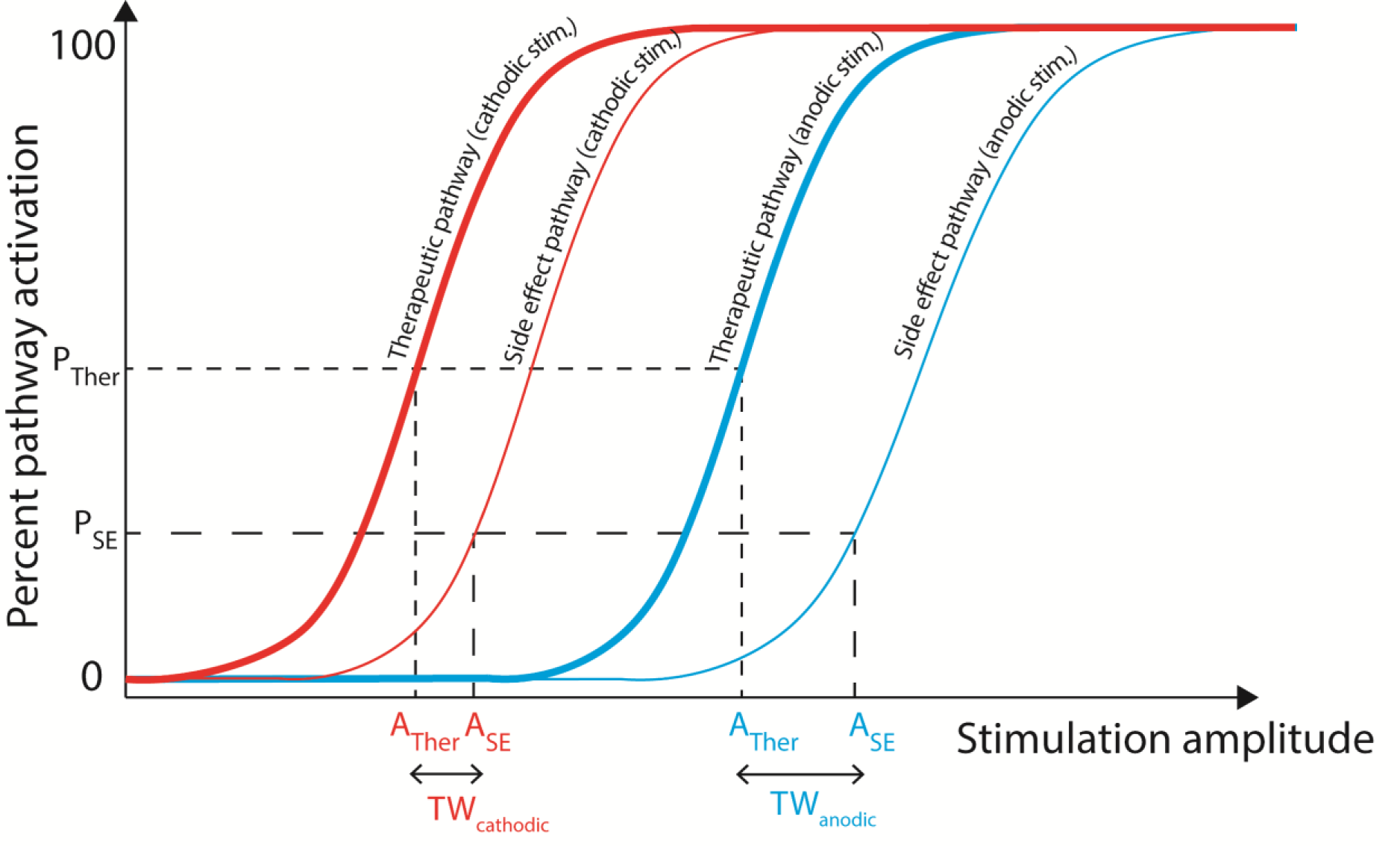
A hypothesized relationship between recruitment curves and clinical thresholds for activation of therapeutic (thick line) and side effect (thin line) pathways in response to cathodic (red) and anodic (blue) stimulation. P = percent; Ther = therapeutic; A = amplitude; TW = therapeutic window; SE = side effect.

Two recent clinical studies have established that monopolar anodic DBS can achieve similar (or even greater) clinical benefit as cathodic, but it requires higher current amplitudes ^6,7^. Side effects were also induced at higher anodic thresholds, but overall the therapeutic window was wider for anodic stimulation. In our experiments, activation of HDP required more anodic current while activation of STN-GP did not, suggesting that HDP may be the pathway responsible for therapeutic benefit observed in clinical studies. If the STN-GP pathway was the main driver of clinical benefit, then we would expect therapeutic thresholds to be similar for anodic and cathodic DBS. CSBT activation required more anodic current in clinical studies which is consistent with our experimental findings. A caveat to this conclusion is that our experiments used low frequency stimulation (10Hz) while clinical DBS employs high frequency (∼130 Hz). A previous study has shown that EP1 amplitude does not depend on cathodic stimulation frequency ^17^, but it remains to be tested whether pathway-specific relative efficiency holds for high frequencies.

Our experiments were not designed to assess therapeutic benefit, but we can compare pathway activation thresholds in our study to threshold amplitudes for therapeutic efficacy in prior studies. We determined that the average cathodic threshold to activate the HDP was 2.46 ± 0.8 mA compared to 5.26 ± 1.4 mA for anodic which is similar to therapeutic benefit amplitudes reported in clinical studies. In Kirsch et al ^6^ therapeutic benefit thresholds were 1.99 ± 1.37 mA for cathodic and 3.36 ± 1.58 mA for anodic, while Soh et al ^7^ reported 3.8 ± 1.6 mA and 4.9 ± 2.1 mA, respectively. In our experiments it took 2.2 times more anodic current than cathodic to achieve the same HDP activation (anodic to cathodic threshold ratio), while in clinical studies the ratio to achieve therapeutic benefit was 1.3-1.7. Overall, our findings suggest that activation of the HDP is likely involved in therapeutic effects of STN DBS, but contribution of other pathways in the target region cannot be ruled out.

### Limitations

Our study has several limitations: 1) the number of stimulation settings examined varied between patients and the recruitment and strength-duration curves had coarse resolution. This was due to variations in the available experimental time in the operating room. Nonetheless, patients showed consistent responses which allowed us to combine results for group analysis; 2) stimulation waveforms had a low-amplitude second recharge phase of opposite polarity as required for safe delivery of electrical stimulation in human subjects ^78^. It is possible that the recharge phase contributed to neural activation although we were careful to exclude stimulation settings where the recharge phase amplitude would be above the activation threshold, and in several patients we utilized an analog stimulation protocol to prolong the second phase for strength-duration curve calculations; 3) DLEP thresholds required for strength-duration curve calculations were difficult to determine (thresholds were consistently below 1mA and recruitment curves noisy in some patients) resulting in fewer data points; 4) we do not know if PD alters pathway excitability and whether our findings are generalizable to heathy state or other conditions where STN DBS is used clinically.

### Conclusions

We provide experimental evidence that relative efficiency of cathodic and anodic stimulation varies for different pathways so that anodic stimulation was the least efficient in activating CSBT, moderately efficient in activating the HDP, and as efficient as cathodic in activating STN-GP pathway. Additionally, we demonstrated that cathodic and anodic stimulation activate the same subcortical pathways in the STN region. Our experiments confirm biophysical model predictions regarding neural activations in the central nervous system and provide evidence that stimulus polarity has differential effects on passing axons, terminal synapses, and local neurons. Comparison of experimental results with clinical DBS studies provides further evidence that the hyperdirect pathway may be involved in the therapeutic mechanisms of DBS.

## Data Availability

The data used in the present study will be made available upon publication of the manuscript in a peer-reviewed journal.

## Acknowledgement

The study was funded by NIH/NINDS (R01 NS125143; K23 NS097576). We thank all the patients for participation.

## Supplementary Materials

**Figure S1.**
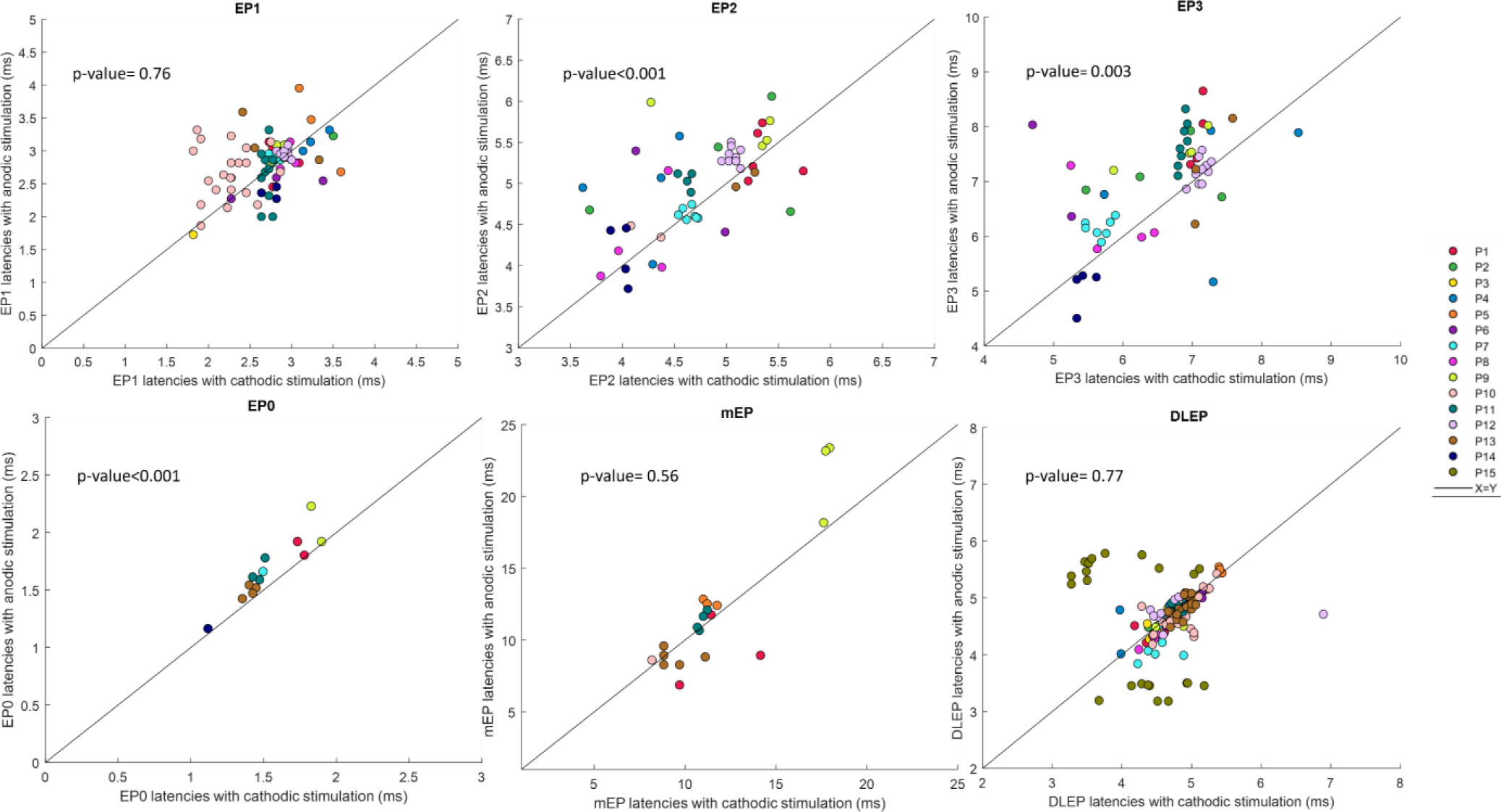
Comparison of evoked potential peak latencies in response to cathodic and anodic stimulation (paired stimulation settings). Each point represents EP response for one stimulation setting pair, colored by patient. Response from one ‘best’ recording channel (largest response) is shown for clarity. The p-values indicates significant differences in EP0, EP2 and EP3 and DLEP pairwise comparisons with most responses lying over the unity (x=y) line indicating delayed EP peaks with anodic stimulation.

**Table S1.** Comparison of average evoked potential amplitudes and latencies in response to cathodic and anodic paired stimulation (all stimulation parameters were the same except stimulus polarity). P-values are from GEE method.

| Mean ± SD | EP1 | EP2 | EP3 | EP0 | mEP | DLEP |
| --- | --- | --- | --- | --- | --- | --- |
| Cathodic amplitude (μV) | 8.506 ± 10.716 | 8.593 ± 11.711 | 11.654 ± 15.200 | 2.800 ± 5.373 | 20.556 ± 42.004 | 27.860 ± 23.149 |
| Anodic amplitude (μV) | 2.300 ± 4.429 | 3.314 ± 6.048 | 4.783 ± 8.435 | 0.315 ± 1.412 | 5.562 ± 16.715 | 26.107 ± 25.764 |
| p-value | <0.001 | <0.001 | <0.001 | 0.005 | 0.004 | 0.63 |
| Cathodic latency (ms) | 2.702 ± 0.388 | 4.729 ± 0.528 | 6.537 ± 0.807 | 1.530 ± 0.220 | 11.669 ± 3.032 | 4.698 ± 0.458 |
| Anodic latency (ms) | 2.801 ± 0.385 | 4.968 ± 0.553 | 6.964 ± 0.922 | 1.665 ± 0.270 | 11.995 ± 4.694 | 4.690 ± 0.516 |
| p-value | 0.76 | <0.001 | 0.003 | <0.001 | 0.56 | 0.77 |

**Figure S2.**
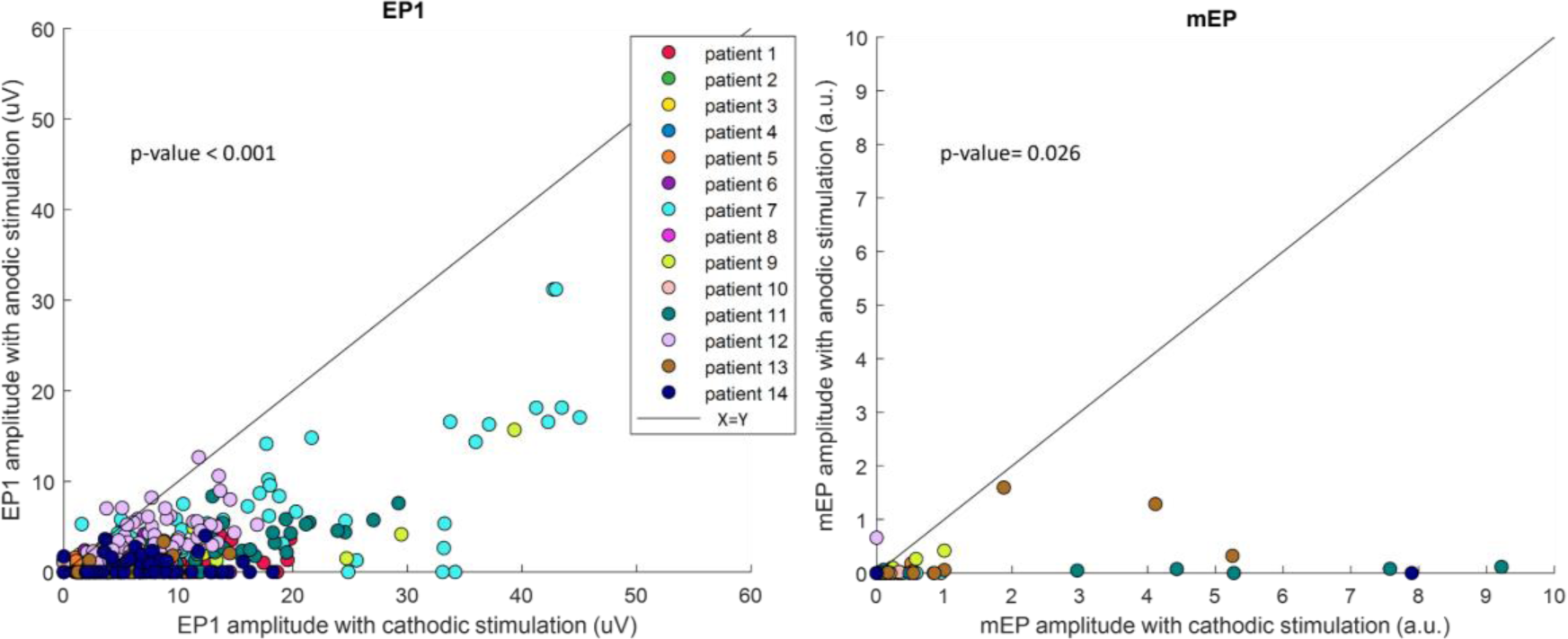
Expanded analysis of EP1 and mEP responses for paired cathodic and anodic stimulation settings. Left: Comparison of cathodic and anodic EP1 amplitudes in all M1 channels. Each point represents EP1 amplitude in response to cathodic its matched anodic stimulation in one M1 channel. Right: Comparison of cathodic and anodic mEP amplitude with average of normalized amplitude over all EMG channels (muscles). Normalized amplitude is sometimes larger than 1, typically for settings with pulse widths longer than 60us because the highest 60 us setting was normalized to 1 for each muscle in each patient.

**Table S2.** Statistical comparison of cathodic and anodic EP1 amplitudes in all M1 channels and by patient. Adjusted p-values are from Bonferroni correction analysis.

|  | Number of<br>M1/total ECoG<br>channels | Number of<br>paired DBS<br>settings | Average EP1<br>amplitude for<br>cathodic<br>stimulation | Average EP1<br>amplitude for<br>cathodic<br>stimulation | Adjusted p-value |
| --- | --- | --- | --- | --- | --- |
| P01 | 9 / 26 | 5 | 10.18 ± 4.89 | 0.80 ± 1.18 | **< 0.001** |
| P02 | 10 / 26 | 4 | 3.79 ± 3.33 | 0.52 ± 0.69 | **0.009** |
| P03 | 11 / 26 | 18 | 0.69 ± 0.95 | 0.05 ± 0.30 | **0.005** |
| P04 | 9 / 26 | 4 | 0.83 ± 1.14 | 0.10 ± 0.33 | **0.027** |
| P05 | 2 / 5 | 6 | 1.08 ± 1.30 | 0.34 ± 0.61 | 2.078 |
| P06 | 1 / 5 | 6 | 2.73 ± 2.82 | 1.38 ± 1.72 | 3.500 |
| P07 | 10 / 26 | 7 | 16.61 ± 12.96 | 6.28 ± 6.61 | **< 0.001** |
| P08 | 3 / 5 | 4 | 5.72 ± 4.66 | 0.08 ± 0.27 | **0.007** |
| P09 | 2 / 5 | 4 | 17.83 ± 12.05 | 3.69 ± 5.16 | 0.109 |
| P10 | 6 / 26 | 24 | 0.41 ± 1.15 | 0.08 ± 0.29 | **0.003** |
| P11 | 7 / 26 | 10 | 10.07 ± 6.80 | 1.64 ± 2.10 | **< 0.001** |
| P12 | 9 / 26 | 10 | 7.72 ± 3.22 | 3.31 ± 2.34 | **< 0.001** |
| P13 | 8 / 26 | 8 | 4.75 ± 3.28 | 0.27 ± 0.67 | **< 0.001** |
| P14 | 7 / 26 | 8 | 7.08 ± 4.78 | 0.82 ± 1.11 | **< 0.001** |

P15 did not have an ECoG electrode.

**Table S3.**
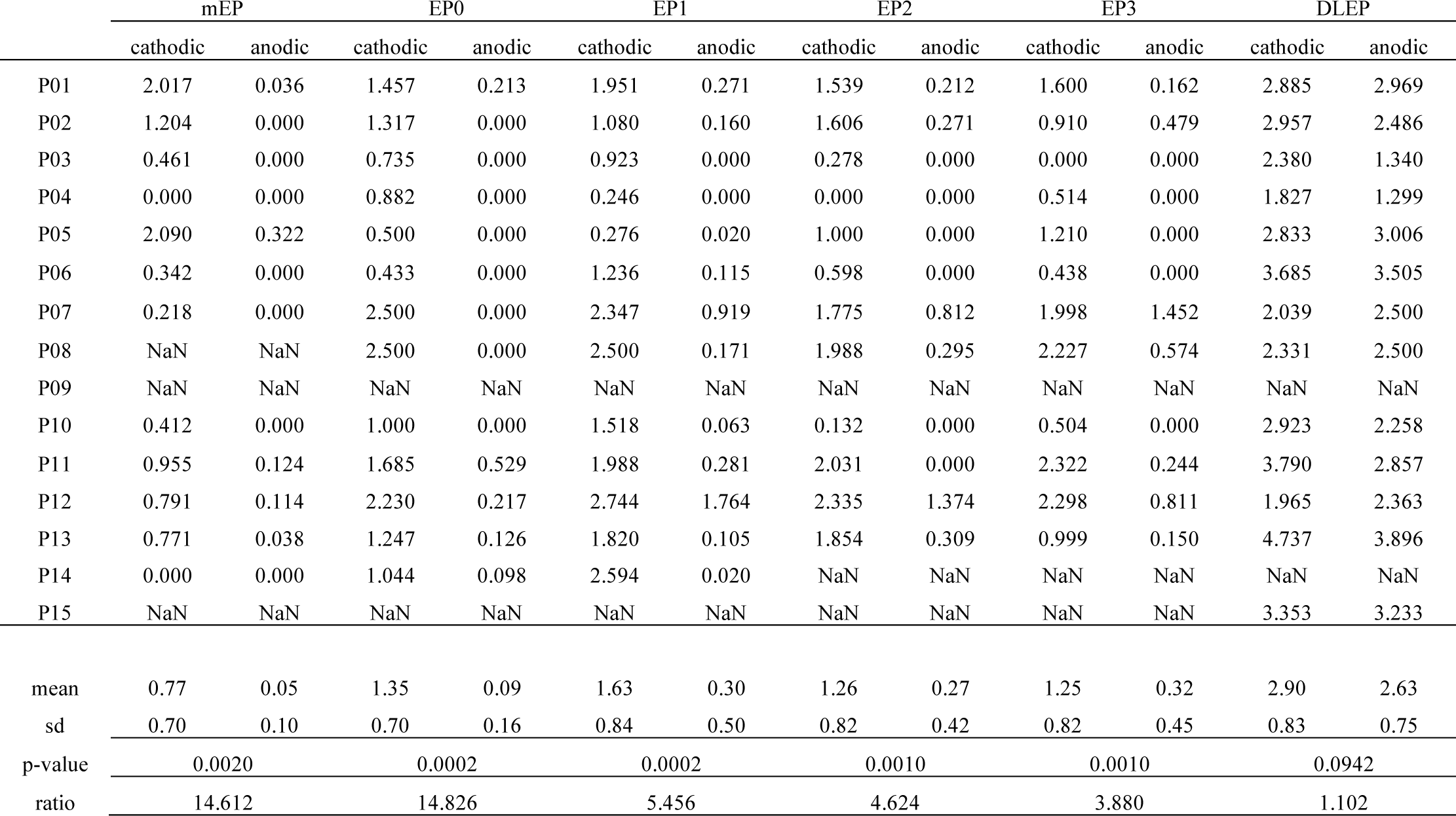
Comparison between area under the curve (AUC) of cathodic and anodic recruitment curves for individual patients.

|  | mEP |  | EP0 |  | EP1 |  | EP2 |  | EP3 |  | DLEP |  |
| --- | --- | --- | --- | --- | --- | --- | --- | --- | --- | --- | --- | --- |
|  | cathodic | anodic | cathodic | anodic | cathodic | anodic | cathodic | anodic | cathodic | anodic | cathodic | anodic |
| P01 | 2.017 | 0.036 | 1.457 | 0.213 | 1.951 | 0.271 | 1.539 | 0.212 | 1.600 | 0.162 | 2.885 | 2.969 |
| P02 | 1.204 | 0.000 | 1.317 | 0.000 | 1.080 | 0.160 | 1.606 | 0.271 | 0.910 | 0.479 | 2.957 | 2.486 |
| P03 | 0.461 | 0.000 | 0.735 | 0.000 | 0.923 | 0.000 | 0.278 | 0.000 | 0.000 | 0.000 | 2.380 | 1.340 |
| P04 | 0.000 | 0.000 | 0.882 | 0.000 | 0.246 | 0.000 | 0.000 | 0.000 | 0.514 | 0.000 | 1.827 | 1.299 |
| P05 | 2.090 | 0.322 | 0.500 | 0.000 | 0.276 | 0.020 | 1.000 | 0.000 | 1.210 | 0.000 | 2.833 | 3.006 |
| P06 | 0.342 | 0.000 | 0.433 | 0.000 | 1.236 | 0.115 | 0.598 | 0.000 | 0.438 | 0.000 | 3.685 | 3.505 |
| P07 | 0.218 | 0.000 | 2.500 | 0.000 | 2.347 | 0.919 | 1.775 | 0.812 | 1.998 | 1.452 | 2.039 | 2.500 |
| P08 | NaN | NaN | 2.500 | 0.000 | 2.500 | 0.171 | 1.988 | 0.295 | 2.227 | 0.574 | 2.331 | 2.500 |
| P09 | NaN | NaN | NaN | NaN | NaN | NaN | NaN | NaN | NaN | NaN | NaN | NaN |
| P10 | 0.412 | 0.000 | 1.000 | 0.000 | 1.518 | 0.063 | 0.132 | 0.000 | 0.504 | 0.000 | 2.923 | 2.258 |
| P11 | 0.955 | 0.124 | 1.685 | 0.529 | 1.988 | 0.281 | 2.031 | 0.000 | 2.322 | 0.244 | 3.790 | 2.857 |
| P12 | 0.791 | 0.114 | 2.230 | 0.217 | 2.744 | 1.764 | 2.335 | 1.374 | 2.298 | 0.811 | 1.965 | 2.363 |
| P13 | 0.771 | 0.038 | 1.247 | 0.126 | 1.820 | 0.105 | 1.854 | 0.309 | 0.999 | 0.150 | 4.737 | 3.896 |
| P14 | 0.000 | 0.000 | 1.044 | 0.098 | 2.594 | 0.020 | NaN | NaN | NaN | NaN | NaN | NaN |
| P15 | NaN | NaN | NaN | NaN | NaN | NaN | NaN | NaN | NaN | NaN | 3.353 | 3.233 |
| mean | 0.77 | 0.05 | 1.35 | 0.09 | 1.63 | 0.30 | 1.26 | 0.27 | 1.25 | 0.32 | 2.90 | 2.63 |
| sd | 0.70 | 0.10 | 0.70 | 0.16 | 0.84 | 0.50 | 0.82 | 0.42 | 0.82 | 0.45 | 0.83 | 0.75 |
| p-value | 0.0020 |  | 0.0002 |  | 0.0002 |  | 0.0010 |  | 0.0010 |  | 0.0942 |  |
| ratio | 14.612 |  | 14.826 |  | 5.456 |  | 4.624 |  | 3.880 |  | 1.102 |  |

**Figure S3.**
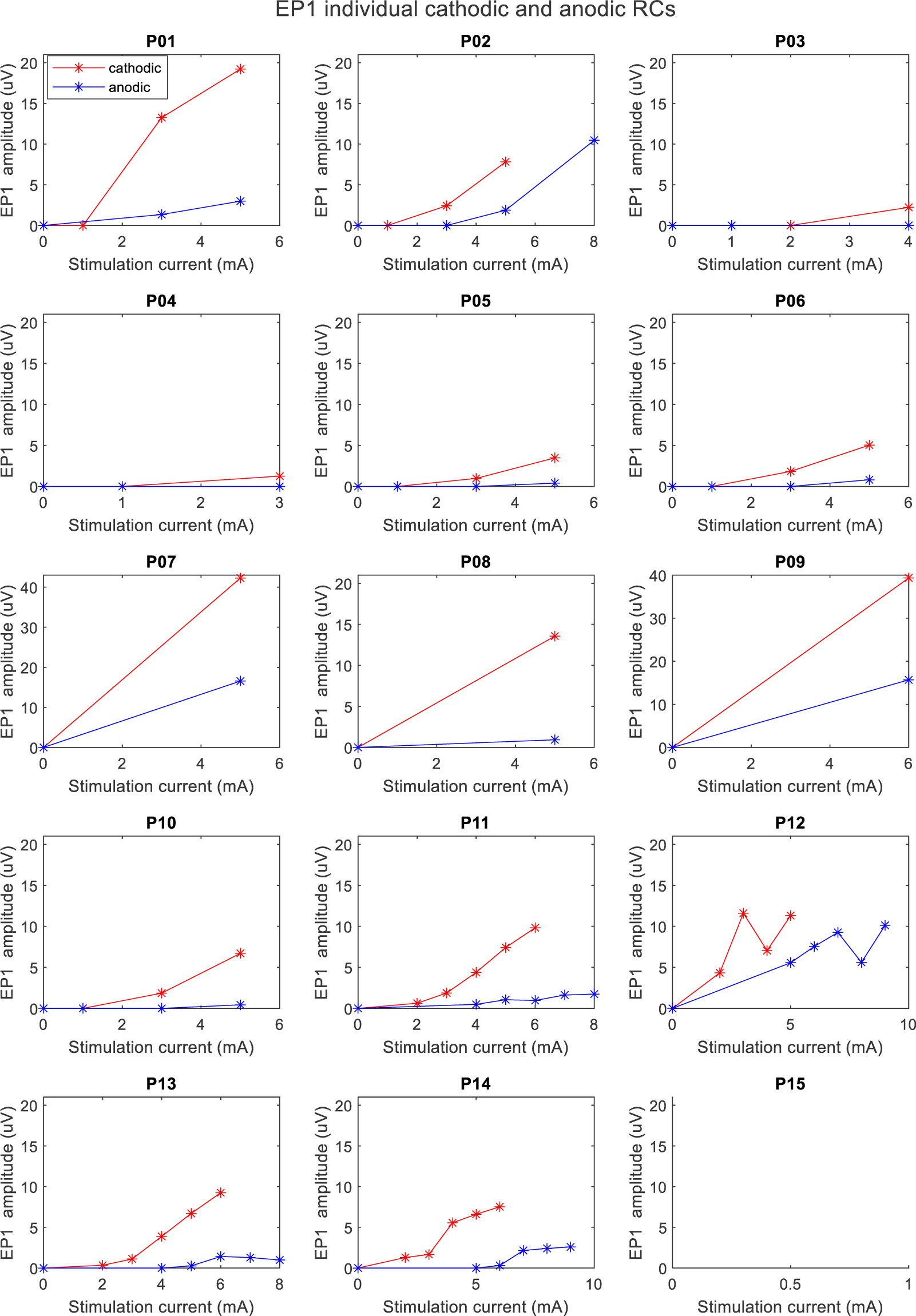
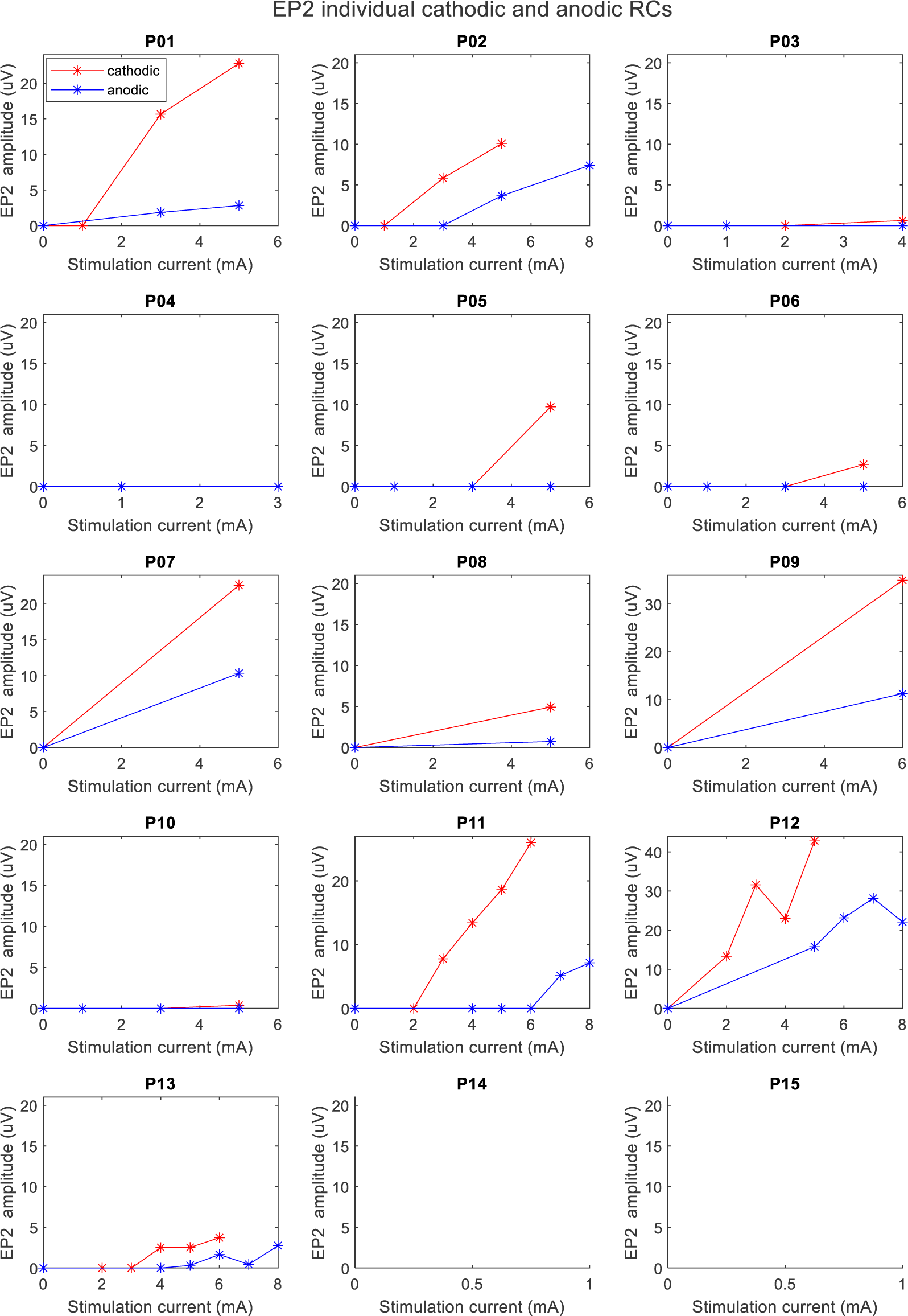
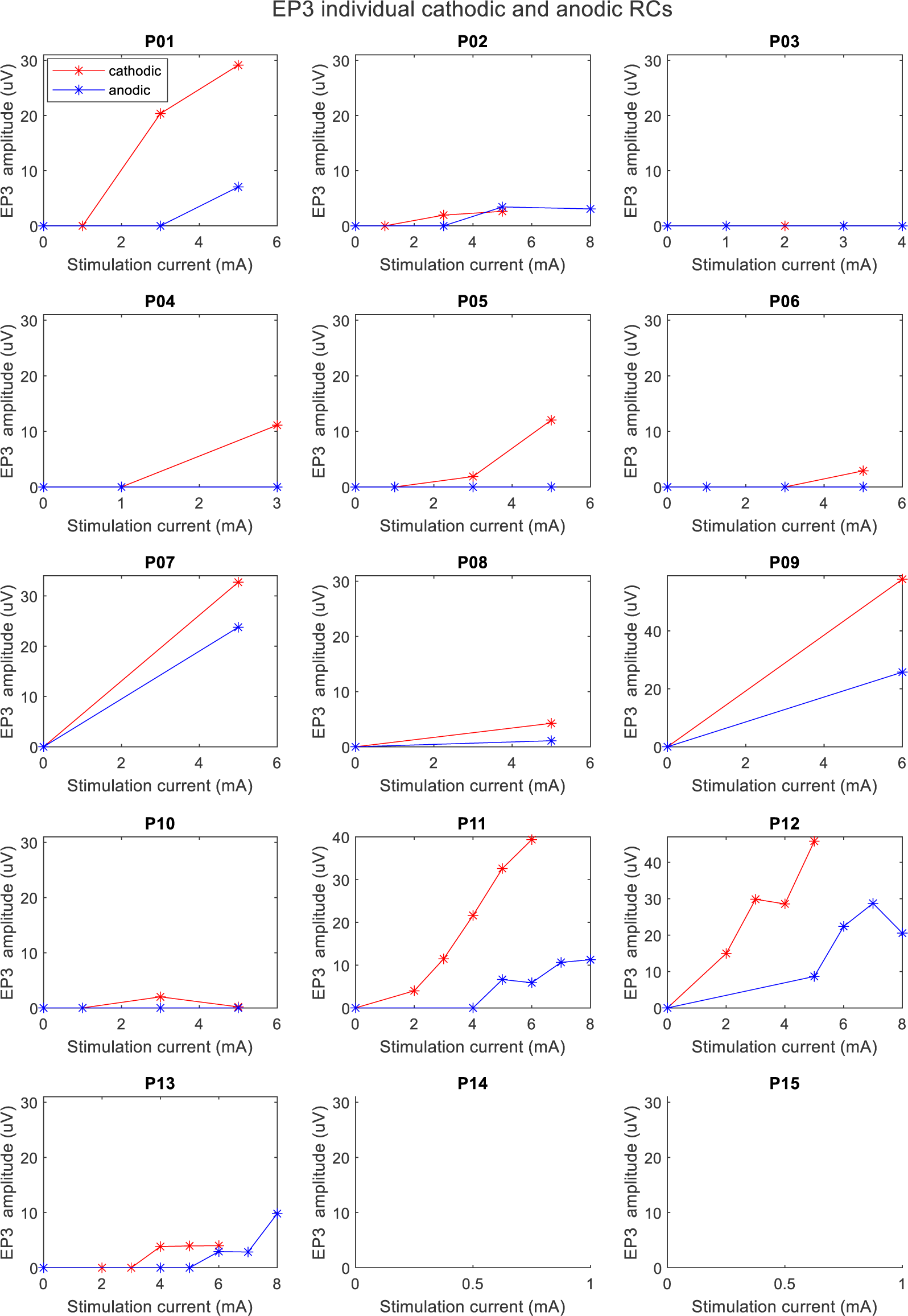
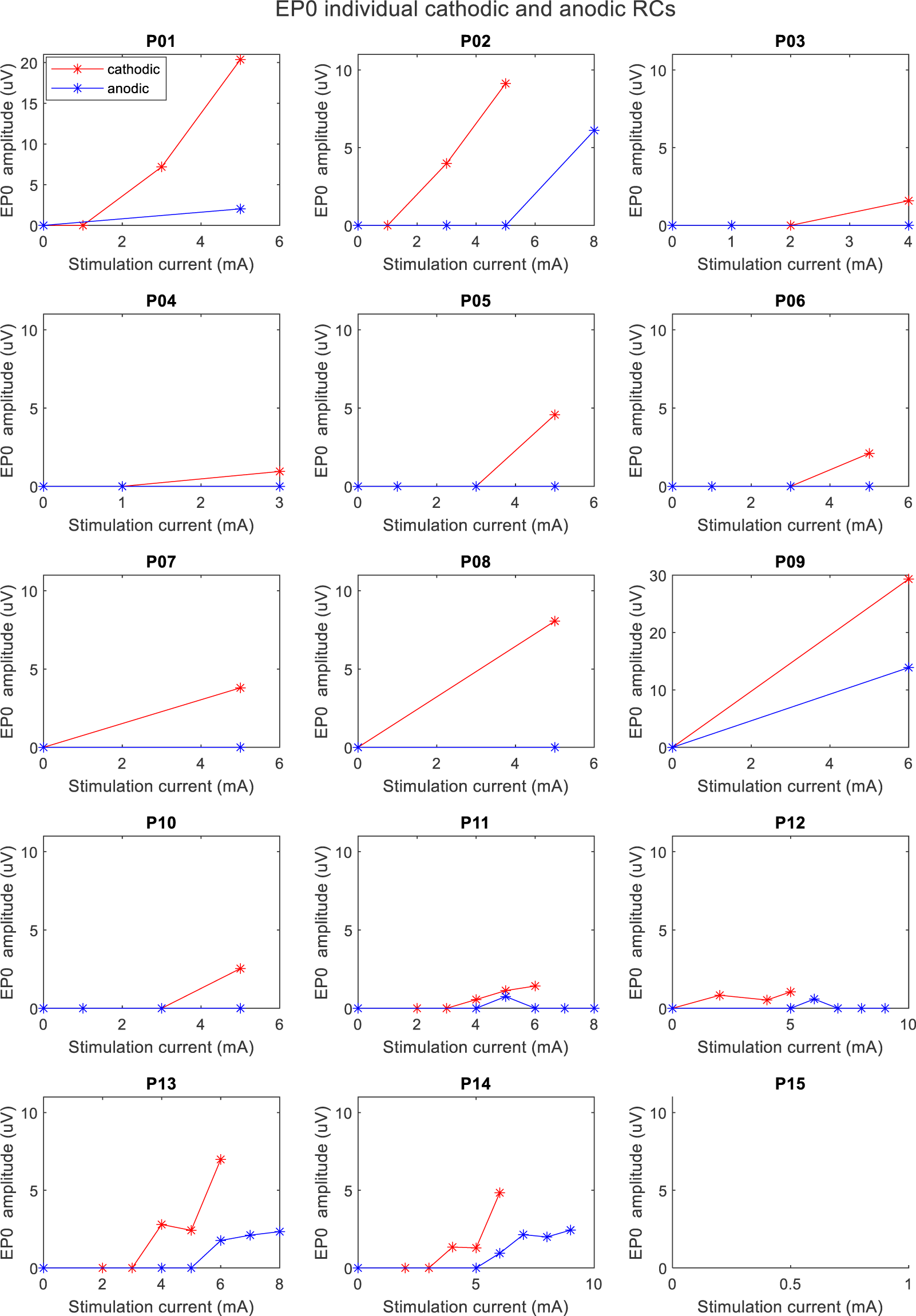
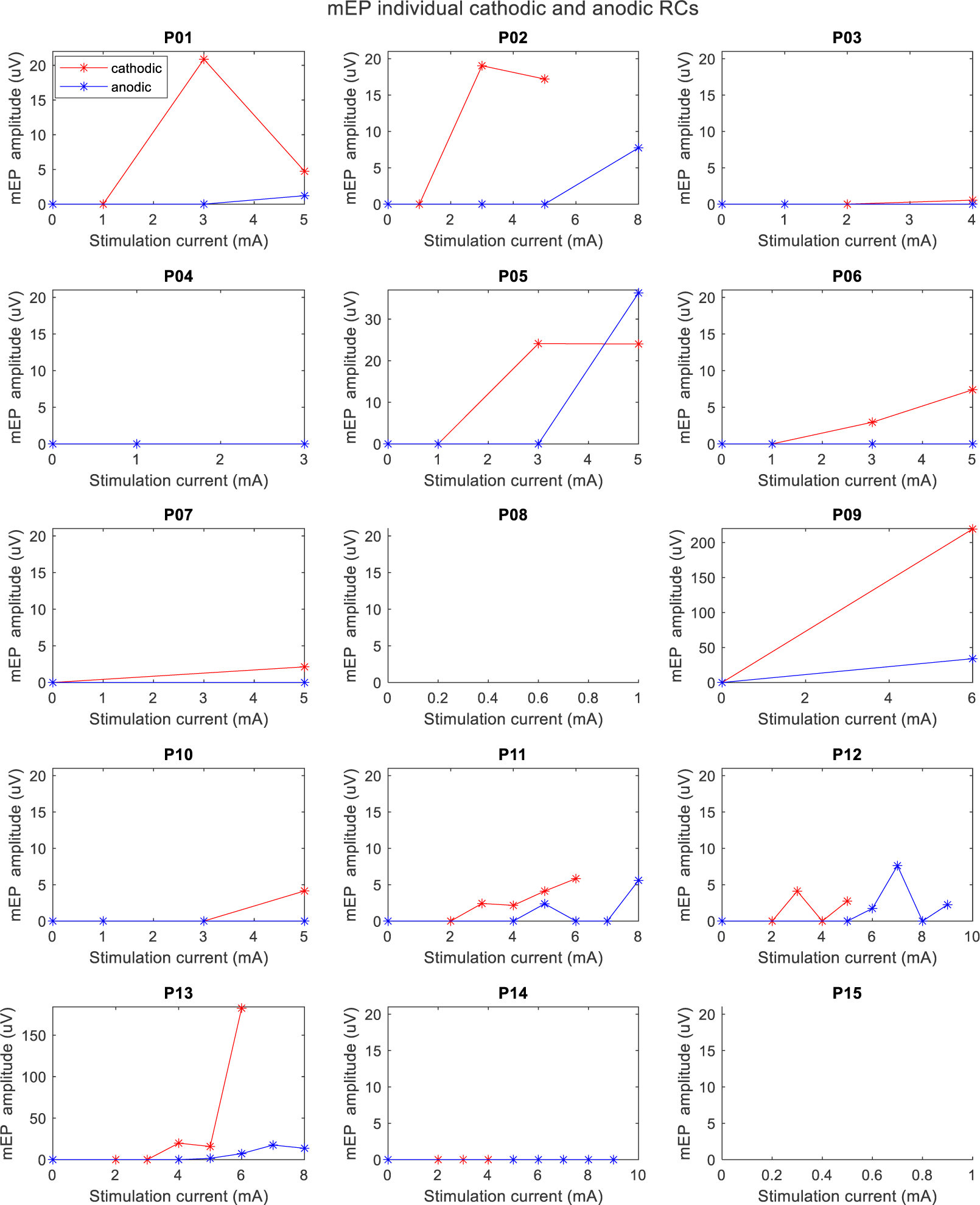
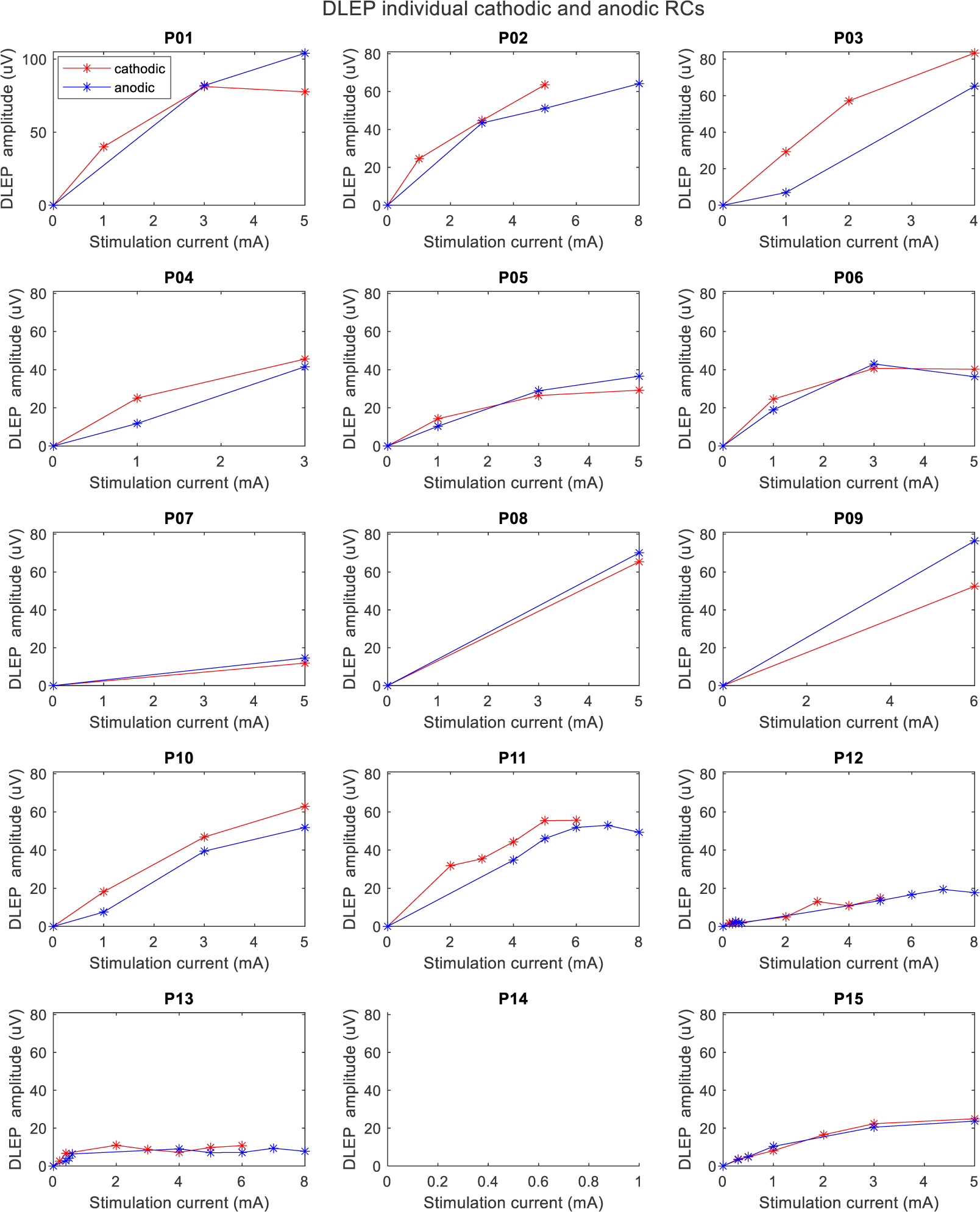
Individual patient recruitment curves for all EP during cathodic (red) or anodic (blue) stimulation. Stimulation was in the bottom (most ventral) DBS contact with 60µs pulse width (100µs for P9). The recording shown is from the ‘best’ channel. For DLEP, stimulation was in the contact that resulted in the largest DLEP response when amplitude-titration data was available for more than one contact.

**Table S4.** Comparison of anodic vs cathodic current thresholds for HDP activation (EP1 amplitude > 1uV)

|  | cathodic (mA) | anodic (mA) | Ratio |
| --- | --- | --- | --- |
| P01 | 1.29 | 3.76 | 2.91 |
| P02 | 2.75 | 5.98 | 2.18 |
| P04 | 1.9 | 3 | 1.58 |
| P11 | 2.3 | 5.6 | 2.43 |
| P12 | 2.16 | 5.20 | 2.41 |
| P13 | 3.69 | 6.69 | 1.81 |
| P14 | 3.14 | 6.58 | 2.10 |
| mean | 2.46 | 5.26 | 2.20 |
| std | 0.80 | 1.40 | 0.44 |

